# CD206^+^IL-4Rα^+^ MACROPHAGES ARE DRIVERS OF ADVERSE CARDIAC REMODELING IN ISCHEMIC CARDIOMYOPATHY

**DOI:** 10.1101/2024.10.01.24314741

**Authors:** Qiongxin Wang, Mohamed Ameen Ismahil, Yujie Zhu, Gregg Rokosh, Tariq Hamid, Guihua Zhou, Steven M. Pogwizd, Sumanth D. Prabhu

## Abstract

**BACKGROUND:** Cardiac CD206^+^ macrophages expand acutely after myocardial infarction (MI) to promote wound healing; however, their role in chronic heart failure (HF) is unknown. We tested the hypothesis that cardiac CD206^+^ macrophages expressing interleukin(IL)-4Rα are key drivers of adverse left ventricular (LV) remodeling in HF.

**METHODS AND RESULTS:** Adult male C57BL/6 mice underwent non-reperfused MI to induce HF, or sham operation, and cardiac macrophages were profiled using flow cytometry. As compared to sham, CD206^+^ macrophages steadily expanded in post-MI hearts during LV remodeling, such that at 8 w post-MI they comprised ∼85% of all macrophages. These macrophages were robustly proliferative and predominantly C-C motif chemokine receptor (CCR2)^-^ and major histocompatibility complex (MHC)II^hi^, with >90% of CD206^+^CCR2^-^ macrophages expressing the resident macrophage marker LYVE-1. CD206^+^ macrophage abundance correlated with LV dysfunction and fibrosis. Nearly half of CD206^+^ macrophages expressed IL-4Rα, and the majority of CD206^+^IL-4Rα^+^ macrophages co-expressed the pro-fibrotic protein found in inflammatory zone (FIZZ)1. IL-4-polarized bone marrow derived CD206^+^ macrophages also exhibited marked upregulation of FIZZ1 and induced FIZZ1-dependent myofibroblast differentiation of cardiac mesenchymal stem cells (cMSCs), in part related to DLL-4/Jagged1-Notch1 signaling. Intramyocardial adoptive transfer of M[IL-4], but not M[IL-10], CD206^+^ macrophages to naïve mice, induced progressive LV remodeling over 4 weeks, increasing fibrosis, cardiomyocyte hypertrophy, and apoptosis. Myeloid-specific IL-4Rα gene deletion in established HF (initiated 4 w post-MI) in IL-4Rα^f/f^LysM-Cre^ERT2^ mice significantly reduced CD206^+^ macrophage proliferation and effectively depleted CD206^+^IL-4Rα^+^ cardiac macrophages. This was associated with abrogation of LV remodeling progression, reduction of cardiac fibrosis, and improved neovascularization. *In vivo* IL-4Rα gene silencing in mice with established HF effectively depleted cardiac CD206^+^IL-4Rα^+^ macrophages and reversed LV remodeling, improving fibrosis, neovascularization, and dysfunction, and suppressed both local and systemic inflammation. Lastly, alternatively activated CD206^+^ and CD163^+^ macrophages were significantly expanded in human failing hearts and correlated with fibrosis. The majority of CD163^+^ macrophages expressed IL-4Rα and FIZZ3, the human homolog of FIZZ1.

**CONCLUSION:** Cardiac CD206^+^IL-4Rα^+^ macrophages proliferate and expand in HF and are key mediators of pathological remodeling and fibrosis, in part through the secretion of FIZZ1. Inhibition of CD206^+^ macrophage IL-4Rα signaling alleviates LV remodeling in ischemic cardiomyopathy.

**CLINICAL PERSPECTIVE:** *What is new?:*

- In the failing heart after myocardial infarction (MI), CD206^+^ macrophages, primarily CCR2^-^, MHCII^hi^, and LYVE1^+^, robustly proliferate with a subpopulation expressing IL-4Rα; myeloid-specific IL-4Rα deletion or IL-4Rα silencing *in vivo* tempered CD206^+^ macrophage proliferation and alleviated LV dysfunction, fibrosis, and remodeling.
- M[IL-4] CD206^+^ bone-marrow derived macrophages induced upregulation of *Fizz1,* and FIZZ1-dependent cardiac mesenchymal myofibroblast differentiation; intramyocardial adoptive transfer of M[IL-4] macrophages induced cardiac remodeling and fibrosis.
- Humans exhibit expansion of alternatively activated macrophages in the failing heart marked by CD206 and CD163 expression, with predominance of a subpopulation expressing IL-4Rα and Fizz3, the human homolog of *Fizz1*.

*What are the clinical implications?:*

- An expanded pool of alternatively activated CD206^+^IL-4Rα^+^ cardiac macrophages are key drivers of LV remodeling in heart failure, inducing an immuno-fibrotic and para-inflammatory response in part dependent on FIZZ1.
- Targeting IL-4 signaling in CD206^+^ macrophages represents a potential therapeutic approach to limit long-term cardiac remodeling in heart failure.

## INTRODUCTION

Heart failure (HF) is characterized by local and systemic inflammation.^1^ In ischemic cardiomyopathy, activated macrophages expand in the failing myocardium and contribute to tissue injury and adverse left ventricular (LV) remodeling.^1–5^ Cardiac macrophages may be classified as C-C motif chemokine receptor (CCR2)^-^ resident macrophages that are embryonically derived and maintained independent of blood monocytes, and CCR2^+^ macrophages that are derived from monocyte recruitment.^6–8^ Acutely after myocardial infarction (MI), resident CCR2^-^ macrophages are lost and replaced by Ly6C^hi^CCR2^+^ monocytes and inflammatory CCR2^+^ macrophages that later transition to reparative macrophages to facilitate wound healing, accompanied by attendant proliferation of surviving resident macrophage populations.^3,6,9–11^ Notably, in chronic ischemic cardiomyopathy, well after MI healing, the expanded cardiac macrophage pool stems from both locally proliferating resident macrophages and recruited CCR2^+^ monocytes,^2,3,5,6,12^ with overall predominance of locally sourced (CCR2^-^) cells.^3,5,12^ CCR2^+^ macrophage abundance correlates with adverse LV remodeling in human HF,^2^ whereas inhibiting CCR2^+^ monocyte infiltration sub-acutely after MI improves LV remodeling in mice,^5^ identifying CCR2 as a potential therapeutic target. In contrast, the roles of locally sourced macrophage populations in the pathogenesis of LV remodeling in established HF are poorly defined.

Resident cardiac macrophages express several marker genes including *Timd4, Lyve1,* and classical alternatively-activated (M2-like) macrophage markers *Cd163* and *Mrc1*.^2,6,13,14^ In the naïve mouse heart, CD206 (*Mrc1*)-expressing resident macrophages are primarily CCR2^-^ and express high levels of the markers *Ym1* and *Fizz1*.^15^ After acute MI, CD206^+^ macrophages expand in the infarct and border zone (peaking at 7 d post-MI)^15,16^ to promote wound healing and limit adverse remodeling.^15,17^ CD206^+^ macrophages are activated by Th2 cytokines IL-4/13, and IL-10.^18–20^ IL-4/13 are important inducers of a reparative and pro-fibrotic phenotype,^18,19,21^ signaling through Type I or II IL-4 receptors containing the common alpha chain IL4Rα, resulting in STAT6-mediated activation of *Arg1*, *Mrc1*, *Fizz1* (*Retnla*), *Ym1*, and secretion of C-C motif chemokine ligands (CCL)-17 and −22.^18,20,22,23^ In vivo administration of IL-4 acutely after MI improves wound healing and remodeling via the modulation of CD206^+^ macrophages. However, this beneficial effect is lost when IL-4 is given late after MI.^15,24^

These prior studies highlight CD206^+^ macrophages as key mediators of the post-MI wound healing response. However, the importance of this macrophage population in the chronically failing heart, which exhibits a considerably different immune cell profile,^1,11^ is unknown. In chronic ischemic cardiomyopathy, locally sourced and proliferating cardiac macrophages exhibit much greater *Mrc1* and *Fizz1* expression than monocyte-derived macrophages.^5^ Moreover, in chronic pulmonary and renal disease, persistent M[IL-4/13] activation or M2-like macrophage recruitment can contribute to tissue fibrosis,^21,25^ whereas we have previously shown that the post-MI failing heart exhibits increased IL-4/13 expression coincident with a Th2 CD4^+^ T-cell profile.^26^ Hence, here we tested the hypothesis that expansion of cardiac CD206^+^ macrophages, activated by IL-4 receptor signaling, are key mediators of LV remodeling in chronic ischemic HF.

## METHODS

### Mouse models

All mouse studies complied with the National Research Council’s Guide for the Care and Use of Laboratory Animals (revised 2011) and were locally approved by Institutional Animal Care and Use Committees at the University of Alabama at Birmingham and Washington University in St. Louis. CD45.2 (#000664) and CD45.1 (#002014) C57BL/6 mice, Fizz*1^-/-^* mice (#029976), and LysM-Cre^ERT^^2^ mice (#031674) were purchased from Jackson Laboratory. Floxed IL4Rα (IL4Rα^f/f^) mice^27^ were originally obtained from Dr. Frank Brombacher (University of Cape Town) and bred with LysM-Cre^ERT2^ mice to generate IL-4Rα^f/f^LysM-Cre^ERT2^ mice that allow for tamoxifen-inducible myeloid-specific deletion of IL-4Rα. To induce Cre-mediated recombination, mice were fed tamoxifen citrate chow (40 mg/kg/d; Inotiv TD.130860).

As 400mg/kg tamoxifen citrate is sufficient to induce recombination,^28^ IL-4Rα^f/f^LysM-Cre^ERT2^ mice were considered to reach stable myeloid-specific IL4Rα deletion after 2 w of feeding. Male mice were used for experiments unless otherwise indicated. Mice were housed in temperature-controlled cages under a 12-hour light-dark cycle and given free access to water and regular chow. A total of 248 male and 37 female mice at 8-10 weeks of age were used. After terminal experiments, mice were euthanized by isoflurane inhalation (2%).

### Murine surgical procedures

Permanent coronary ligation to induce large MI and ischemic HF, and control sham surgery, were performed under general anesthesia with 1-1.5% inhaled isoflurane and mechanical ventilation as previously described.^4,26,29–31^ For macrophage adoptive transfer, cell suspensions (1×10^6^ polarized macrophages in 50 μL sterile PBS) or sterile PBS control alone were injected intramyocardially into the free wall of left ventricle (10 μL/site x 5 sites) in naïve C57BL/6 male recipient mice.

### Echocardiography

M-mode and 2-dimensional echocardiography were performed under 2% inhaled isoflurane anesthesia (with 98% O2) and continuous ECG monitoring using a VisualSonics Vevo 3100 High-Resolution System and 30 MHz MX400 transducer with a heated, bench-mounted adjustable rail system.^4,26,29,32,33^ Imaging was performed in the parasternal long-axis and short-axis views. LV chamber volume in end-diastole (EDV) and end-systole (ESV) were quantitated using the area-length method from long-axis images captured in ECG-gated Kilohertz Visualization mode. Systolic function was indexed by LV ejection fraction (EF), calculated as [(EDV−ESV)/EDV] * 100. Stroke volume was defined as EDV−ESV. LV mass (mg) was calculated as 1.05*[(IVSWTd+EDD+PWTd)^3^-(EDD)^3^], where IVSWTd and PWTd are interventricular septal and LV posterior wall thickness at end-diastole, and EDD is the LV end-diastolic diameter derived from M-mode tracings.

### Gene expression analysis

Total RNA from LV remote zones (≈10 mg), macrophages and cMSCs was extracted using Trizol (Invitrogen, 15596026) and Qiagen RNAeasy Columns (QIAGEN, 74204)^34^ and reverse transcribed to cDNA using the High-Capacity cDNA Reverse Transcription Kit (Applied Biosystems, 43-688-14). cDNAs were subjected to qPCR using SYBR Green Master Mix (Applied Biosystems, A25742) and run on a ViiA 7 RT PCR System (Applied Biosystems).^32,35–37^ Forward and reverse primer sequences are listed in *Supplemental Table 1*.

### Multiplex Luminex assay

Peripheral blood was coagulated on ice and centrifuged to separate serum. Cytokine Luminex assay of serum samples was performed in 96-well Bio-Plex Pro Mouse Cytokine microplates and analyzed using the Luminex reader (BIO-RAD, Bio-PlexTM 200 system) according to the manufacturer’s protocol.^34^ Bio-Plex Pro Mouse Cytokine 23-plex Assay (M60009RDPD), MIP-2 Set (171G6006M), TGF-β1 Set (171V4001M), Standards Group I (171I50001), Group II Standards (171I60001), TGF-β Standard (171X40001) and Reagent Kit V (12002798) were all purchased from BIO-RAD.

### Immunoblot analysis

Immunoblotting was conducted using standard protocols as described previously,^33,34,36^ using antibodies against mouse Notch1 (Cell Signaling, 4380), NICD (Cell Signaling, 4147) and α-SMA (Abcam, ab7817). GAPDH (Cell Signaling, 97166) and α-tubulin (Santa Cruz, 5286) were used as loading controls.

### Immune cell isolation and flow cytometry

Live mononuclear cells were isolated from peripheral blood and heart tissue and processed for flow cytometry as previously described.^26,29,38^ After 1% paraformaldehyde fixation, isolated cells were resuspended in MACS buffer (PBS with 2 mM EDTA and 0.5% BSA) and preincubated for 10 min at 4°C with anti-mouse CD16/CD32 antibody (eBioscience, #14-0161-82) to block Fcγ receptors.^32^ Cells were then stained for surface membrane markers upon incubation with fluorophore-labeled anti-mouse CD45, F4/80, MERTK, CD11b, Ly6G, Ly6C, CD3, CD4, CD8, IL-4Rα, IL-10Rα, MHCII, LYVE1, and CCR2, and, for human cardiac mononuclear cells, anti-human CD45, CD64, CD206 and CCR2 for 1 h. Mouse cardiac mononuclear cells were then permeabilized with 0.5% Tween-20 for 20 minutes, followed by staining with anti-mouse CD206 and Ki67 for 1 h. All staining was performed on ice to maintain cell morphology. Flow cytometry was performed using a FACSdiva LSR-II flow cytometer (BD) and analyzed using FlowJo software v10.8.0. Specific details regarding the fluorophore-conjugated antibodies used are listed in *Supplemental Table 2*.

### (Immuno)histological analysis

Formalin-fixed, paraffin-embedded heart tissues were sectioned at 5 μm thickness, deparaffinized, and rehydrated. Cardiomyocyte cell membranes and capillaries were stained using Alexa Fluor (AF)555-conjugated wheat-germ agglutinin (Invitrogen, #W32464) and AF488-conjugated isolectin-IB4 (Invitrogen, #121411), respectively.^4,26,29,32^. After antigen retrieval, LV sections were stained with anti-mouse CD206 (1:50, R&D AF2535, secondary anti-goat AF555), Ki67 (1:50, eBioscience 14-5698-82, secondary anti-rat AF488), CD45.2 (1:50, eBioscience 14-0454-82, secondary anti-mouse AF647), α-actinin (1:100, Sigma A2172, secondary anti-mouse AF555), IL-4Rα (1:50, Invitrogen PA5-103142, secondary anti-rabbit AFF647) and FIZZ1 (1:50, R&D MAB-1523, secondary anti-rat AF488) or anti-human CD163 (1:100, Leica Biosystems CD163-L-CE, secondary anti-mouse IgG1 AF647), IL-4Rα (1:50, R&D MAB230, secondary anti-mouse IgG2a AF488) and FIZZ3 (1:50, R&D AF1359, secondary anti-goat AF555) antibodies. After permeabilization with 20 μg/ml proteinase K, α-actinin stained LV sections were incubated with TUNEL reaction mix (Biotium 30063) for 2h at 37°C. CD206 or CD163 stained LV sections were incubated with 15 μM heat-dissociated collagen hybridizing peptide 5-FAM Conjugate (F-CHP) (3Hekix FLU300) overnight at 4°C. In addition, LV frozen sections (8 μm) were fixed with acetone/chloroform, rehydrated, and stained with anti-mouse F4/80 (1:100, Abcam ab6640, secondary anti-rat AF488) and α-actinin (1:100, Sigma A2172, secondary anti-mouse AF555) antibodies. Nuclei were stained with DAPI. Images were acquired using a Nikon A1 confocal microscope. Fibrosis was measured in LV sections using Masson’s Trichrome staining, and images were captured by a Nikon 80i Nomarski DIC microscope. Collagen (blue) staining percent of total cross-sectional area was determined as previously described.^4,26,29,32^ Staining was quantified from more than 5 high-power fields (40x) per section using Nikon NIS-Elements software, version 4.60.00.

### Bone marrow macrophage culture and adoptive cell transfer

C57BL/6 (CD45.2) or *Fizz1*^-/-^ mouse bone marrow (BM) cells were harvested aseptically and cultured in RPMI 1640 medium, supplemented with 10% fetal bovine serum (FBS), 1% penicillin-streptomycin (PS) and 30 ng/mL recombinant mouse macrophage-colony-stimulating factor (M-CSF, R&D, 416-ML) at 37°C. The supernatant was replaced with fresh complete medium containing 30 ng/mL M-CSF every 3 days for the attached cells, following the protocol of Trouplin et al.^39^ After 7 days of differentiation, BM-derived macrophages (BMDMs) were cultured in RPMI 1640 with 10% FBS and 1% PS for 24 h for M0 (resting) macrophages, with 40 ng/mL recombinant mouse IL-4 (R&D, 404-ML) for M[IL-4] macrophages, and with 40 ng/mL recombinant mouse IL-10 (R&D, 417-ML) for M[IL-10] macrophages. Polarized macrophages were then mechanically detached, and cell aliquots were stained with anti-mouse CD45, F4/80 and CD206 (*Supplemental Table 2*) and analyzed using a flow cytometer to evaluate macrophage purity. After 24 h of polarization, M0, M[IL-4] and M[IL-10] cells were rinsed with PBS and cultured in FBS-free medium for 3 h. The macrophage conditioned medium was subsequently collected for treatment of cardiac mesenchymal stem cells and fibroblasts, as described below. The polarized macrophages were used for studies of intramyocardial adoptive cell transfer into naïve male CD45.1 C57BL/6 mice as described above.

### *In vivo* antisense oligonucleotide administration

Second-generation chimeric 20-base IL-4Rα antisense oligonucleotides (ASOs) (CCGCTGTTCTCAGGTGACAT) and control mismatched oligonucleotides (MM oligos) (CCGCTCATCACTGCTGACAT) were synthesized by Integrated DNA Technologies (IDT). All 20-base oligonucleotides were designed to avoid murine immune-stimulatory motifs, and contained phosphorothioate backbones and 2’-O-methoxyethylribose (MOE)-modification on bases 1–5 and 16–20 as described previously.^40,41^ Oligonucleotides were suspended in sterile saline and administered every 3 days (40 mg/kg, total volume 10 μL saline/g body weight) by intraperitoneal injection.^41^ Male C57BL/6 sham-operated and HF mice (4 w post-MI) with comparable degrees of LV remodeling were randomized to receive either IL-4Rα ASOs (Ref 219428745, IDT) or MM oligos (Ref 219428746, IDT) for 4 w.

### *In vivo* phagocytosis assay

To assess cardiac macrophage phagocytosis *in vivo*, pHrodo™ green-E. coli bioparticles (Invitrogen, P35366) were injected intraperitoneally to mice (200 μg/mouse) 4 h before sacrifice. After sacrifice, heart mononuclear cells were collected and cell surface marker staining performed with anti-mouse CD45, F4/80, MERTK and CD206 as described above. Bioparticle uptake in live macrophages was measured by flow cytometry.

### Cardiac mesenchymal stem cell (cMSC) studies *in vitro*

Sca1^+^CD90^+^CD31^-^DDR2^-^ cMSCs were isolated from mouse hearts and maintained in supplemented [DMEM]/Ham’s F-12 culture medium as previously described,^33^ and used within 3-5 cell passages for all experiments. For isolated cell stimulation studies, cMSCs were treated at the indicated times with serum-free conditioned RPMI 1640 medium from M0, M[IL-4], or M[IL-10] macrophages described above (or control media alone), with some experiments performed in the presence of either 1.5 μg/mL Fizz1 neutralization antibody or IgG isotype control (antibodies-online.com, ABIN636644 and ABIN1881001, respectively). In separate studies, cMSCs were treated with recombinant mouse Fizz1 (Leinco Technologies, R1140) at the indicated concentrations and times in serum-free RPMI 1640 medium.

### Collagen gel-contraction assays

These were performed as previously described.^33^ Briefly, cMSCs in ice-cold 10% FBS DMEM medium mixed with 1 mg/mL rat tail collagen I were plated in 48-well tissue culture plates at a density of 3×10^5^ cells/well. The cell suspension was incubated at 37°C for 30 minutes until gelation. After gel formation, an additional 0.5 mL of 10% FBS DMEM medium was added to each well and incubated overnight at 37°C in a 5% CO2 incubator. The supernatant was then removed and the gels treated with 0.5 mL conditioned RPMI 1640 medium from M0, M[IL-4], or M[IL-10] macrophages with 10% FBS (or media alone control), with or without anti-Fizz1 or IgG isotype control (1.5 μg/mL), as described above. Some experiments were performed upon incubation with IL-4 or IL-10 alone (40 ng/mL) in 10% FBS RPMI 1640 medium. The cells were cultured and treated for 5 d with medium change every 2 d. On day 5, the gels were detached from the wells using a pipette tip and incubated overnight. On day 6, the gels were imaged with a digital camera and gel area was measured using Image J software (1.51j8, NIH).

### siRNA transfection of cMSCs

cMSCs were transfected with siRNA using INTERFERin Reagent (Polyplus, 101000028) according to the manufacturer’s protocol. Control siRNA (4390843) and siRNA targeting Notch1 (4390771) were purchased from Invitrogen.

### Cardiac fibroblast studies *in vitro*

Mouse cardiac fibroblasts were purchased from iXCells biotechnologies (10MU-015) and grown in fibroblast growth medium (MD-0011) at 37°C in a humidified 5% CO2 atmosphere. Cells were used within 1-3 passages and treated with M0, M[IL-4], or M[IL-10] conditioned medium for 24 h.

### Human studies

Following patient consent, failing heart tissue was procured from patients with advanced (NYHA IV) HF with reduced EF undergoing LV assist device (LVAD) placement at the University of Alabama at Birmingham (UAB) under the auspices of Institutional Review Board (IRB) approved protocol #X090810003. Non-failing control heart tissues were obtained from donor hearts technically unsuitable for heart transplantation from the Alabama Organ Center. All protocols using human heart samples were approved by the UAB IRB. Failing and control human heart tissues were used for cardiac immune cell isolation and subsequent flow cytometry, as well for histological analysis. Clinical characteristics of human HF subjects and donor controls are shown in *Supplemental Table 3*.

### Statistical Analysis

All group data are mean ± SD. All statistical analyses were performed with GraphPad Prism 9 (GraphPad Software). Normality was assessed using the Shapiro-Wilk test, whereas group variances were compared using the Brown-Forsythe test. For normally distributed data, a two-tailed unpaired Student’s *t* test with equal or unequal variance was used for two-group analyses, and 1- or 2-way ANOVA with Tukey’s post-test was used for multi-group analyses. Additionally, 2-way repeated-measures ANOVA with Dunnett’s multiple comparison test was used to analyze time series pre-post data. For non-normal distributions, the Mann-Whitney U test was used for 2-group analyses. Pearson R correlation test was used for linear dependence analyses. Specific approaches are in the figure legends. A *p* value of <0.05 was considered statistically significant.

## RESULTS

### Cardiac CD206^+^ macrophages progressively expand during the development of chronic HF post-MI

Cardiac macrophage subsets were profiled using flow cytometry at various timepoints following non-reperfused large MI during the development of ischemic HF,^4,26,29,36^ and in sham-operated controls (*Figure 1A*). As shown in *Figures 1B-C and S1A*, following MI there was significant cardiac accumulation of both overall CD45^+^ leukocytes and total F4/80^+^MERTK^+^ macrophages in a biphasic manner, with high levels at 48 h after injury, subsequent decline by 2 w, followed by a second phase of re-expansion up to 8 w post-MI during chronic LV remodeling and failure. In the failing heart 8 w post-MI, macrophages comprised nearly two-thirds (∼63%, or ∼140,000 cells per heart) of all cardiac leukocytes and were increased by ∼9-fold over macrophage levels in sham-operated hearts. Similar dynamics were observed for the blood levels of pro-inflammatory Ly6C^high^ monocytes, an important source of infiltrating macrophages,^3,5,10,32,42^ during the progression of ischemic HF. No significant differences were observed in the levels of patrolling Ly6C^low^ monocytes between sham-operated and HF mice at any time point evaluated (*Figures S2A-C*).

**Figure 1.**
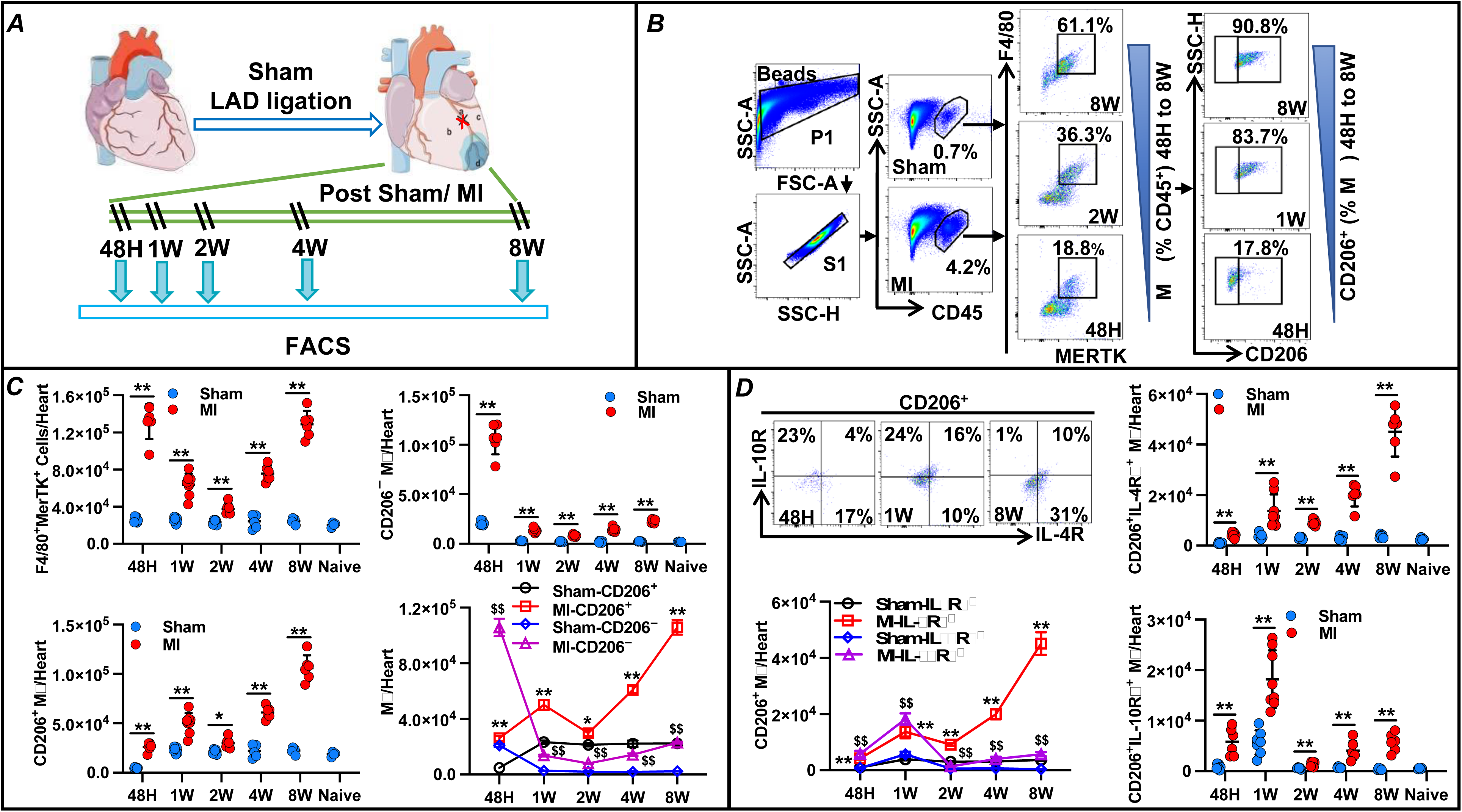
Cardiac CD206^+^ macrophages robustly expand and express IL-4Rα in heart failure (HF). ***A***, Protocol for cardiac macrophage profiling by FACS in C57BL/6 mice at the indicated time points after non-reperfused myocardial infarction (MI) or sham operation. ***B*** and ***C***, representative flow cytometry scatter plots (***B***) and group quantitation for cardiac macrophages (F4/80^+^MerTK^+^), and CD206^+^ and CD206^−^ subpopulations (***C***) from sham and MI mice at the indicated times post-operation. Quantitation from naïve mice also shown. ***D***, Representative flow plots and group data for IL-4Rα and IL-10Rα expression in cardiac CD206^+^ macrophages from the same mouse groups. Statistics: unpaired Student’s t test; \*\**p*<0.01 vs respective sham group in ***C****-**D***; ^$$^*p*<0.01 vs Sham-CD206^−^ in ***C***, ^$$^*p*<0.01 vs Sham-IL-10Rα^+^ in ***D***. N=6-8 per group.

We next further evaluated macrophage subsets in the mouse heart. Naïve murine hearts harbored low numbers of macrophages (∼20,000/heart), of which ∼90% expressed the resident macrophage marker CD206 (*Figure 1C*). However, acutely (48 h) after MI, CD206^+^ macrophages comprised only ∼20% of overall macrophages in the heart, in the setting of a marked expansion of CD206**^−^** macrophages early after MI. Thereafter, both the frequency and absolute number of CD206^+^ macrophages steadily increased in post-MI hearts during the progression of LV remodeling, such that at 8 w post-MI ∼85% of cardiac macrophages expressed CD206. In contrast, CD206**^−^** macrophage levels decreased markedly by 2 w post-MI, and then exhibited smaller sustained increases in chronically failing hearts thereafter. Both populations were significantly increased as compared to sham-operated hearts at all timepoints post-MI. Hence, macrophages expand significantly in the chronically remodeled and failing heart post-MI, and most of these macrophages express CD206.

### Cardiac CD206^+^ macrophages robustly express IL-4Rα in chronic HF

Two key activators of CD206^+^ macrophages are IL-4/13, acting via Type I or II IL-4 receptors (IL-4R) containing the common subunit IL4Rα,^43–45^ and IL-10, signaling through the IL-10 receptor (IL-10R) complex upon binding to the high affinity IL-10Rα (IL-R1) subunit.^46,47^ As illustrated in *Figure 1D*, both CD206^+^IL4Rα^+^ and CD206**^+^**IL-10Rα^+^ macrophage subpopulations were significantly increased in comparison to sham-operated hearts at all time points after MI. However, there was substantial expansion of the CD206^+^IL4Rα^+^ population over time, such that at 8 w post-MI, levels of CD206**^+^**IL-4Rα^+^ cells (∼45,000 per heart) were ∼8-fold higher than CD206**^+^**IL-10Rα^+^ cells and comprised ∼42% of all CD206^+^ macrophages in the chronically failing heart. In contrast, at 1 w post-MI, during the healing phase, CD206**^+^**IL-10Rα^+^ macrophages were more abundant than CD206**^+^**IL-4Rα^+^ cells, comprising ∼40% of all CD206^+^ macrophages in the heart. Notably, neither IL-4Rα nor IL-10Rα were abundantly expressed by cardiac CD206^−^ macrophages (<3% of cells) at any timepoint post-MI, and CD206**^−^** macrophages expressing either IL-4Rα or IL-10Rα numbered fewer than 500 cells/heart in the chronic stages of LV remodeling (*Figure S1B*). In addition, IL-4Rα and IL-10Rα expression was also very low in blood neutrophils (<1.5%), Ly6C^low^ monocytes (<2%) and Ly6C^high^ monocytes (<5%) in both sham and HF mice (*Figure S2*).

Regarding Th2 cytokines, serum levels of IL4 and IL-10 were significantly higher in MI mice as compared with sham mice during both the healing phase (1 w) and chronic remodeling phase (8 w) post-MI, whereas levels of IL-13 and TGF-β1 were comparable in sham and MI mice at all timepoints (*Figure S3A*). In contrast, serum levels of the pro-inflammatory cytokines TNF-α, IFN-γ, IL-1β, MCP-1 and MIP-2α in HF mice were increased over sham controls during both the early and late phases of remodeling after MI (*Figure S3B*), consistent with ongoing systemic inflammation in HF mice. Hence, CD206^+^ macrophages in the failing heart primarily express IL-4Rα (versus IL-10Rα); this is accompanied by increased circulating levels of both IL-4 and IL-10.

### CD206^+^ macrophages in the failing heart proliferate, are primarily CCR2^-^, MHCII^hi^, and LYVE1^+^ and correlate with LV dysfunction and fibrosis

At 8 w post-MI, HF mice exhibited significant LV systolic dysfunction and fibrosis (*Figure 2A and S4A*). Cardiac CD206^+^ macrophage abundance inversely correlated with LVEF (r = 0.695), and directly correlated with border and remote zone fibrosis (r = 0.755 and 0.644, respectively). Tissue co-staining with anti-CD206 and fluorescein-conjugated collagen hybridizing peptide (CHP) that recognizes denatured collagen^48^ revealed that CD206^+^ macrophages were primarily found in areas of collagen turnover in the failing heart, with >98% of macrophages associated with CHP-positive regions in the infarct and border zones, and >80% CHP-associated cells in the remote myocardium (*Figure 2B*). To infer the source of CD206^+^ macrophage expansion in the failing heart, we measured the expression of markers CCR2, MHCII, and LYVE1 by flow cytometry (*Figure 2C*). In sham hearts, >90% CD206^+^ macrophages were CCR2^-^, and of these, ∼95% were LYVE-1^+^, indicating a primarily resident macrophage population. In failing hearts, the proportion of CD206^+^CCR2^-^LYVE1^+^ macrophages decreased significantly but remained high at ∼80% of total CD206^+^ cells, suggesting resident cell predominance but with an increased contribution from monocyte-derived macrophages. Moreover, in both sham and failing hearts, the majority (∼70%) of CD206^+^ macrophages were MHCII^hi^.

**Figure 2.**
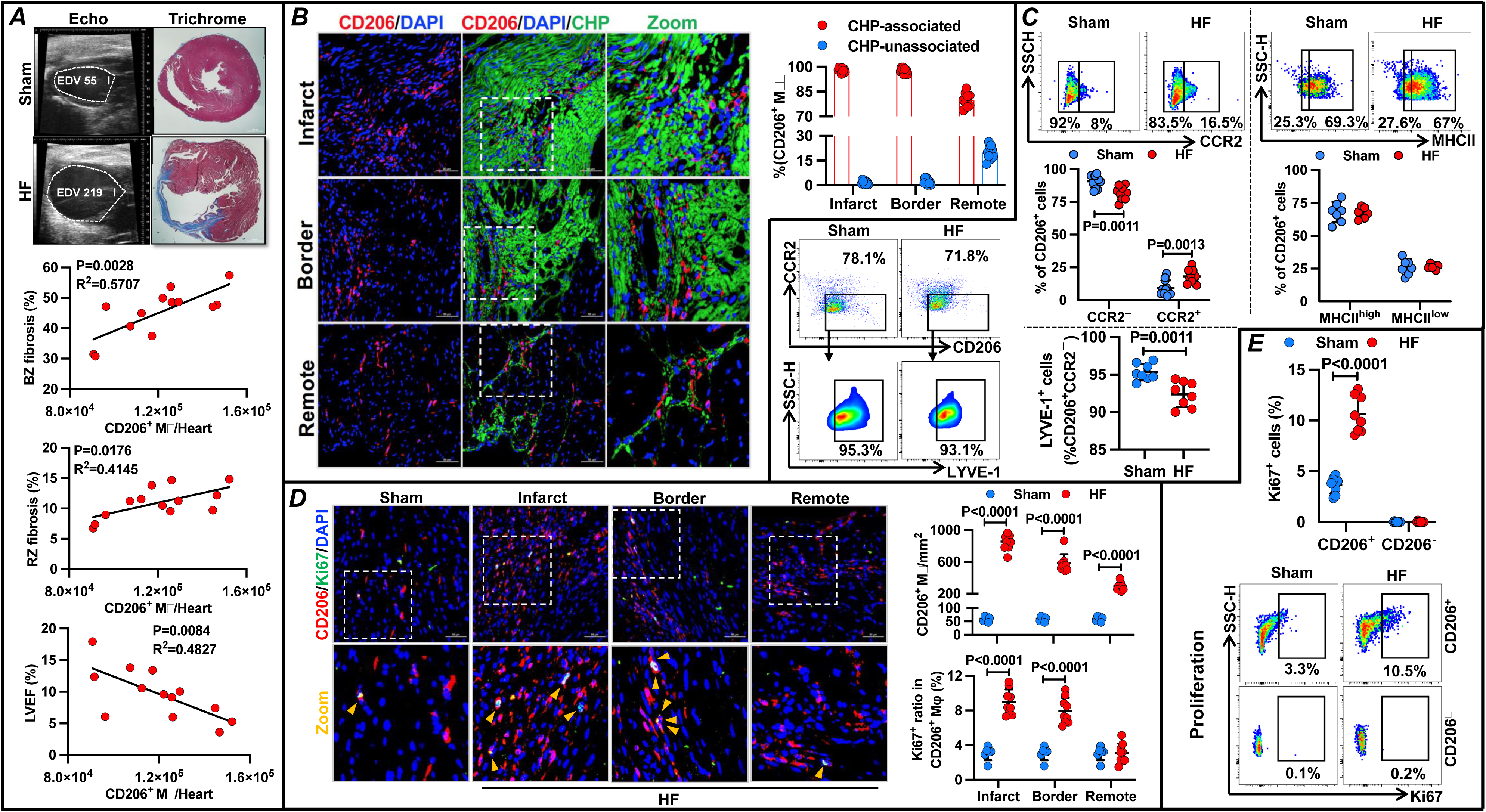
Cardiac CD206^+^ macrophages are proliferative, primarily MHCII^hi^ and LYVE1^+^, and correlate with LV dysfunction and fibrosis in heart failure (HF). *A*, Representative long-axis 2-dimensional echocardiograms at end-diastole and heart trichrome stains from HF mice 8 w after non-reperfused myocardial infarction (MI) and sham controls, along with correlations between cardiac CD206^+^ macrophage number and LV ejection fraction (LVEF), and border and remote zone (BZ and RZ) fibrosis in failing hearts. *B*, Representative low and high magnification confocal images of failing heart (8 w post-MI) infarct, border, and remote zones (immuno)stained for CD206 (red) and collagen hybridizing peptide (CHP) (green), with quantitation of CHP-associated and -unassociated CD206^+^ macrophages. DAPI (blue) was used to label nuclei. Scale bar 50 μm. *C*, Flow plots and quantitation of CCR2 and LYVE1 expression in CD206^+^ macrophages and LYVE-1 expression in CD206^+^CCR2^−^ macrophages in sham and HF hearts 8 w post-surgery, *D*, Low and high magnification confocal images of CD206 (red) and Ki67 (green) immunostains of sham, and the infarct, border and remote zones of failing hearts 8 w post-surgery, and quantitation of CD206^+^ macrophages and Ki67 expression frequency. Nuclei labeled by DAPI (blue); yellow arrows indicate CD206^+^Ki67^+^ cells. Scale bar 50 μm. *E*, Flow plots and quantitation of Ki67 expression in both CD206^+^ and CD206^−^ cardiac macrophages from the same groups. Statistics: Pearson R correlation test in *Aa*; unpaired Student’s t test in *C*-*E*. N=4-13 per group as indicated.

Resident macrophage expansion requires local cell proliferation. As shown in *Figure 2D and S4B*, immunostaining for CD206 and the cell proliferation marker Ki67 revealed that both total CD206^+^ and Ki67^+^CD206^+^ macrophages were robustly expanded in the failing heart (8 w post-MI) as compared to sham. Cell density was greatest in the infarct zone (855 cells/mm^2^; 9% Ki67^+^), followed by the border zone (584 cells/mm^2^; ∼8% Ki67^+^) and then the remote zone (290 cells/mm^2^, Ki67 positivity ∼3% similar to sham hearts). Complementary assessment using flow cytometry demonstrated that in both sham and failing hearts only CD206^+^ macrophages exhibited Ki67 positivity (*Figure 2E*). Moreover, as compared to sham, failing hearts evidenced ∼3-fold higher levels of CD206^+^Ki67^+^ macrophages (∼10% total), confirming augmented local proliferation of CD206^+^ macrophages. In parallel, myocardial gene expression of Th2 cytokines IL-4 and IL-10 (but not IL-13) and classical alternatively-activated macrophage markers *Mrc1 (CD206), Arg1, Ym1* and *Fizz1* was significantly increased in failing versus sham hearts (*Figure S4C*), Concomitantly, the expression of fibrotic genes *(Col)1α1, Col1α2*, and *Col3α1* was markedly increased in failing hearts. Collectively, these data support expansion of a Th2/M2-like immuno-fibrotic response in the chronically failing heart post-MI.

### IL-4 polarization *in vitro* induces a profibrotic phenotype in CD206^+^ macrophages

Bone marrow-derived macrophages (BMDMs) were polarized with either IL-4 or IL-10 (40ng/mL), or medium alone (M0 control), for 24 h (*Figure 3A*). Flow cytometry indicated that while BMDMs were primarily CD206^+^ under resting (M0) conditions, both IL-4 and Il-10 exposure augmented BMDM CD206 expression to ∼98-99% of total cells. As shown in *Figure 3B*, the expression of signature M2 genes^15,18,20,49,50^ *Mrc1, Arg1*, and *Ym1* significantly increased in both M[IL-4] and M[IL-10] macrophages (with much more *Arg1* and *Ym1* expression in M[IL-4] cells) as compared to non-polarized M0 macrophages. In contrast, expression of *Fizz1* (found in inflammatory zone 1), a protein secreted during Th2 responses^51,52^ and implicated in myofibroblast differentiation and lung fibrosis,^53,54^ only increased in M[IL-4] macrophages, and to extraordinary levels (>7,500-fold). Conversely, *B7-H4* was exclusively, albeit modestly, upregulated in M[IL-10] macrophages. Changes in macrophage gene expression were noted after 4 h of polarization (*Figure S5A*) and were maintained or augmented after 48 h (*Figure S5B*). A range of responses was observed for inflammatory and repair-associated genes in BMDMs after 24 h of polarization *in vitro* (*Figure S6A*). Surprisingly, there was no upregulation of profibrotic genes TGF-β1 and platelet-derived growth factor (PDGF)-BB in M[IL-4] and M[IL10] macrophages as compared to M0 cells, although there was augmented expression of tissue inhibitor of matrix metalloproteinase (TIMP)-1 and TIMP-3 in both M[IL-4] and M[IL-10] macrophages.

**Figure 3.**
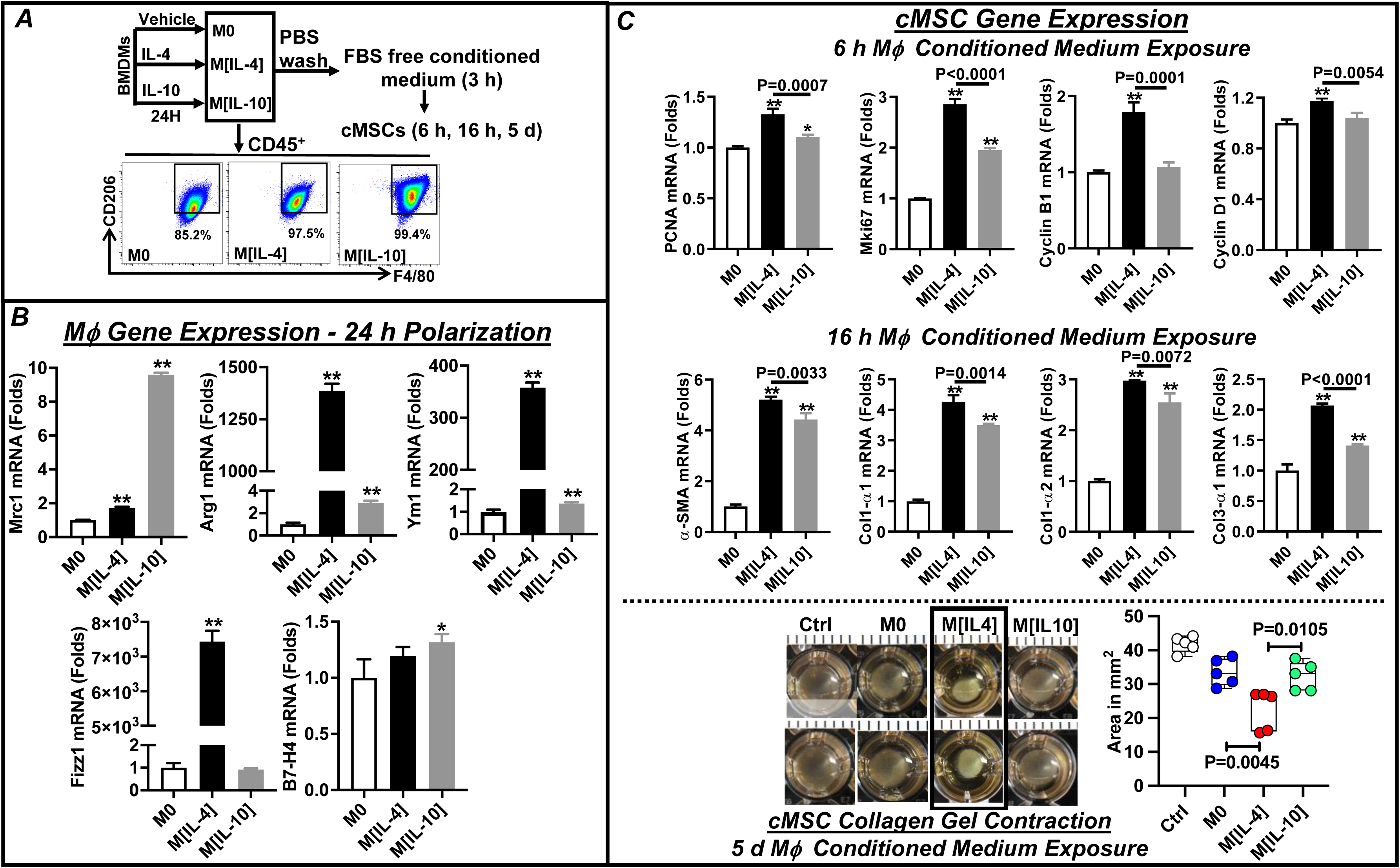
IL-4-polarized bone marrow derived macrophages (BMDMs) induce cardiac mesenchymal stem cell (cMSC) myofibroblast differentiation *in vitro*. *A*, Protocol for BMDM polarization and cMSC treatment with macrophage conditioned medium, and representative flow plots of F4/80 and CD206 expression in polarized BMDMs. *B*, Expression of the indicated genes in M0, M[IL-4], and M[IL-10] macrophages 24 h after polarization. *C*, Expression of the indicated genes in cMSCs treated with M0, M[IL-4], and M[IL-10] conditioned media for the indicated times, and representative cMSC-imbued collagen gel contraction assays upon exposure to M0, M[IL-4], and M[IL-10] conditioned media, or media alone (Ctrl), and quantitation of gel areas. Statistics: 1-way ANOVA with post-hoc Tukey’s multiple comparison test; * *p*<0.05, ** *p*<0.01 vs. M0. N=5-6/group.

In parallel experiments, conditioned medium was collected from M0, M[IL-4] and M[IL-10] macrophages and applied to Sca1^+^CD90^+^CD31**^−^**DDR2**^−^** cardiac MSCs^33^ (*Figure 3A*). cMSC gene expression was then evaluated after 6 and 16 h of exposure. Short-term (6 h) exposure to M[IL-4] medium induced significant upregulation of the proliferation genes *PCNA*, *Mki67, CyclinB1* and *CyclinD1* as compared to M0-medium, and to a greater extent than upon exposure to M[IL-10] medium. Longer-term (16 h) exposure induced similar expression responses for cMSC pro-fibrotic genes, including α-smooth muscle actin *(α-SMA*), *Col1α1, Col1α2*, and *Col3α1* (*Figure 3C*). The functional impact of macrophage polarization on cMSCs was assessed using collagen gel contraction assays^33^ after 5 d of exposure to M0, M[IL-4], M[IL-10] medium, or medium alone control. As illustrated in *Figure 3D*, M[IL-4] medium exposure induced robust gel contraction, significantly greater than with exposure to either M0 or M[IL-10] medium (or control), consistent with vigorous cMSC myofibroblast differentiation. Importantly, exposure to complete medium containing IL-4 or IL-10 alone (40 ng/mL) did not induce cMSC gel contraction (*Figure S6B*), indicating an indispensable role for CD206^+^ macrophages in IL-4-induced cMSC myofibroblast differentiation. Similarly, 24 h treatment with M[1L-4] medium robustly increased α-SMA protein expression in mouse cardiac fibroblasts as compared with similar treatment with M0 or M[IL-10] medium (*Figure S6C*). Collectively, these data suggest that IL-4-mediated macrophage polarization supports pro-fibrotic responses in concert with striking upregulation of *Fizz1*.

### Adoptive transfer of M[IL-4] macrophages to naïve hearts induces cardiac dysfunction and fibrosis

To evaluate the in vivo cardiac effects of M[IL-4] and M[IL-10] macrophages, we adoptively transferred 1×10^6^ M0, M[IL-4], or M[IL-10] bone marrow-derived macrophages from CD45.2 male mice to naïve WT CD45.1 male mice (PBS control) via intramyocardial injection, with subsequent follow-up over 4 w (*Figure 4A*). Immunostaining of injection sites of recipient hearts 30 minutes post-injection confirmed successful transfer of M0, M[IL-4], and M[IL-10] donor macrophages (*Figure S7A*). Serial echocardiography of recipient mice revealed progressive and significant LV chamber dilatation, with increased EDV and ESV, and systolic dysfunction, with reduced LVEF, along with LV hypertrophy, evidenced by increased LV mass, IVSWTd, and PWTd by echocardiography and increased normalized heart weight, in M[IL-4] macrophage recipients, but not in PBS, or M0 or M[IL-10] macrophage recipients (*Figures 4B and S7B and D*). Stroke volume was comparable over time in all four groups (*Figure S7C*). Histological analysis showed significantly increased patchy interstitial fibrosis in M[IL-4]-recipient hearts as compared with PBS, M0, and M[IL-10] recipient hearts (*Figure 4C*), along with augmented cardiomyocyte cross-sectional area and apoptosis (*Figures 4D and S7E*). Importantly, CD45.2 and CD206 co-staining of recipient hearts demonstrated donor macrophage persistence in adoptive transfer recipients, but to a significantly greater level in M[IL-4] recipient mice. M[IL-4] (but not M0 or M[IL-10]) macrophage transfer also induced expansion of native resident CD206^+^ macrophages. Both CD45.2^+^ donor and CD45.2^−^ recipient CD206^+^ macrophages expansion occurred primarily at sites of collagen turnover and fibrosis in recipient hearts, with stronger such association for donor macrophages (*Figures 4E and S7F*), There was no evidence of pro-inflammatory Ly6C^hi^ monocytosis in any recipient mouse group (*Figure S7G*), suggesting the absence of systemic inflammation after transfer. Collectively, these data establish that M[IL-4] polarized macrophages are sufficient to induce chronic cardiac remodeling, hypertrophy, and fibrosis, and a microenvironment supporting CD206^+^ macrophage expansion.

**Figure 4.**
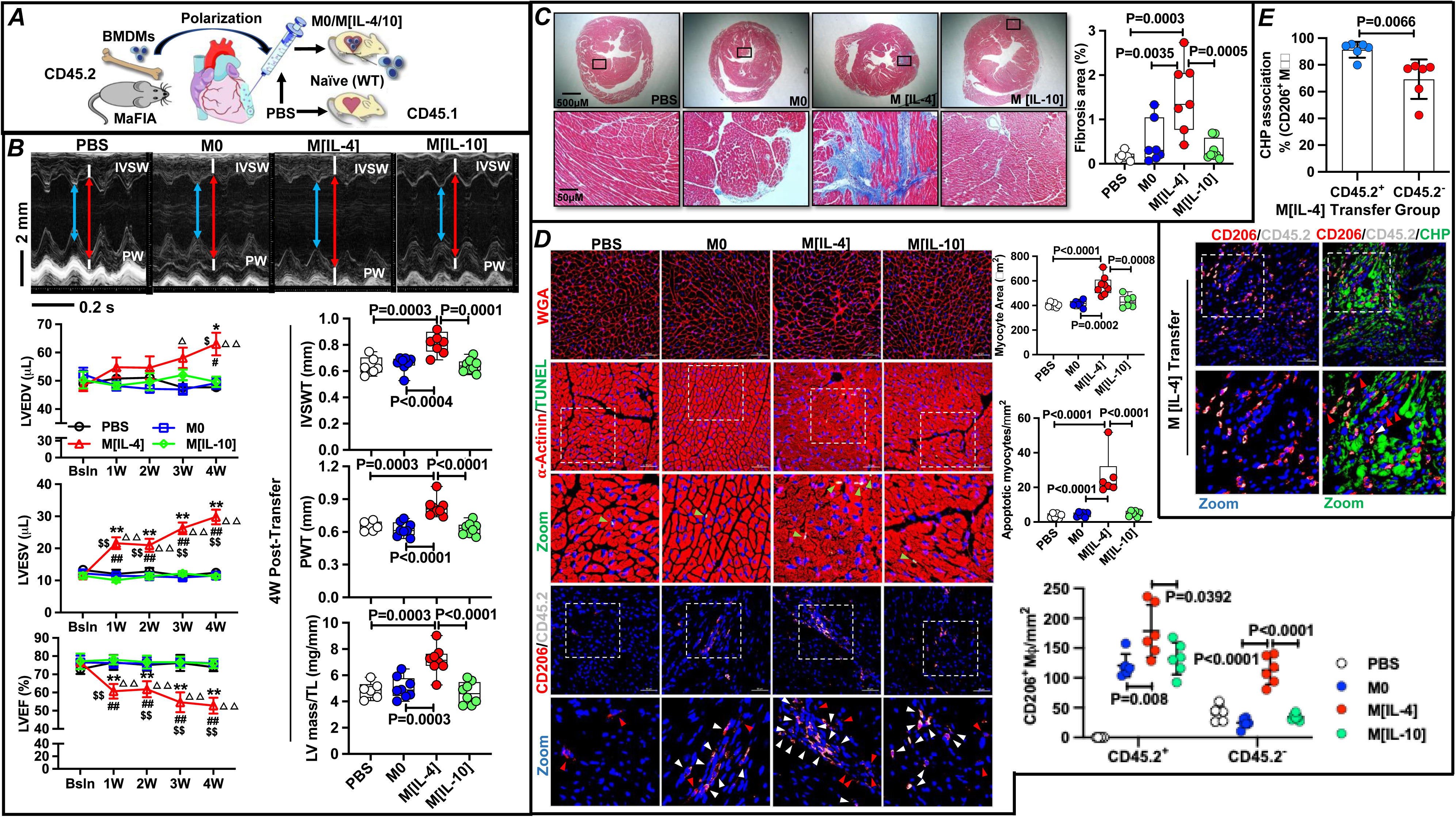
Adoptive transfer of M[IL-4] macrophages to naïve hearts induces cardiac dysfunction and fibrosis. ***A***, Protocol for intramyocardial adoptive transfer of M0, M[IL-4], and M[IL-10] bone marrow derived macrophages (BMDMs) or PBS vehicle to naïve mice. ***B***, M-mode echocardiograms of M0, M[IL-4], and M[IL-10] BMDM recipient mice 4 w post-transfer (PBS control), along with group data for LV end-systolic and end-diastolic volume (ESV and EDV), ejection fraction (EF) at indicated times post-transfer, and for echocardiographic LV mass normalized to tibia length (TL), and end-diastolic interventricular septal and posterior wall thickness (IVSWT and PWT) at 4 w post-transfer. ***C***, Representative trichrome stains of recipient hearts at 4 w post-transfer and corresponding quantitation of cardiac fibrosis. ***D***, *Top*, cardiac wheat-germ agglutinin (WGA, red) staining and quantitation of myocyte cross-sectional area; *Middle*, low and high magnification images of TUNEL (green) stained nuclei in cardiomyocytes (α-Actinin, Red) and quantitation of apoptotic myocyte nuclei; and *Bottom*, Low and high magnification confocal images of CD206 (red) and CD45.2 (gray) immunostains and quantitation of CD206^+^CD45.2^+^ and CD206^+^CD45.2^−^ macrophages in recipient hearts, all at 4 w post-transfer. Green arrows indicate apoptotic myocyte nuclei, red arrows indicate CD206^+^CD45.2^−^ cells, and white arrows indicate CD206^+^CD45.2^+^ cells. DAPI (blue) was used to label nuclei. ***E***, Low and high magnification confocal images of CD206 (red) and CD45.2 (gray) immunostaining and CHP staining (green), and quantitation of CHP-associated and - unassociated, CD45.2^+^ and CD45.2^−^ CD206^+^ macrophage ratios in M[IL-4] recipient hearts 4 w post-transfer. Nuclei were labeled by DAPI (blue). Red arrows indicate CHP-unassociated CD206^+^CD45.2^−^ cells and white arrows indicate CHP-unassociated CD206^+^CD45.2^+^ cells. Statistics: repeated-measures 1-way ANOVA with post-hoc Dunnett’s multiple comparison test for time series within groups in ***B***, 1-way ANOVA with post-hoc Tukey’s multiple comparison test between groups in ***B-D***, and unpaired Student’s t test in ***E***; \**p*<0.05, \*\**p*<0.01 M[IL-4] vs. PBS; ^#^*p*<0.05, ^##^*p*<0.01 M[IL-4] vs. M0; ^$^*p*<0.05, ^$$^*p*<0.01 M[IL-4] vs. M[IL-10]; **^Δ^***p*<0.05, **^ΔΔ^***p*<0.01 vs. Bsln. N=6-8 per group as indicated.

### Myeloid-specific IL-4Rα deletion in HF suppresses cardiac CD206^+^ macrophage expansion and improves LV remodeling

Among myeloid cells, IL-4Rα expressions occurred primarily in macrophages, with low levels in monocytes and neutrophils (*Figure S2*), and among macrophages, primarily in CD206^+^ macrophages (*Figures 1D and S1B*). Hence, to establish the role of CD206^+^IL4Rα^+^ macrophages in HF, we used IL-4Rα^f/f^LysM-Cre^ERT2^ mice. Given the importance of the IL-4 axis in wound healing after acute MI^15,24^ and our observations that cardiac CD206^+^IL-4Rα^+^ macrophage expansion in ischemic HF occurs most prominently >4 w post-MI, we induced myeloid IL-4Rα deletion during late remodeling. Male and female IL-4Rα^f/f^ and IL-4Rα^f/f^LysM-Cre^ERT2^ HF mice with comparable LV remodeling at 4 w post-MI (or sham-operated controls) were randomized to receive tamoxifen diet for 6 weeks (*Figure 5A*). In male mice, echocardiography revealed significant LV dilation and systolic dysfunction in both HF groups 4 w post-MI as compared to sham (*Figure 5B*). In contrast, as compared with tamoxifen-fed IL-4Rα^f/f^ HF mice, tamoxifen treatment for 6 w in IL-4Rα^f/f^LysM-Cre^ERT2^ HF mice resulted in lower LVEDV and ESV, better LVEF and stroke volume (SV), and smaller heart size and normalized heart and lung weight (*Figures 5B, S8A-B*). Moreover, histological analysis revealed less border and remote zone fibrosis, smaller cardiomyocyte area, and improved capillary density in male IL-4Rα^f/f^LysM-Cre^ERT2^ HF mice versus IL-4Rα^f/f^ HF mice (*Figures 5C, S8C*). Myeloid IL-4Rα deletion in HF in male mice also decreased levels of total leukocytes, CD206^+^ and CD206^−^ (and CD206^−^IL4Rα^+^) macrophages, CD206^+^IL4Rα^+^ and CD206^+^IL10Rα^+^ macrophages, and proliferating Ki67^+^CD206^+^ macrophages in the heart, as well as Ly6C^hi^ and Ly6C^hi^IL-4Rα^+^ monocytes (but not Ly6C^low^ monocytes) in the blood, as compared to HF mice without myeloid IL-4Rα deletion (*Figures 5D-F, S8D-F*). There were no effects of myeloid IL-4Rα deletion on any parameter in sham male mice.

**Figure 5.**
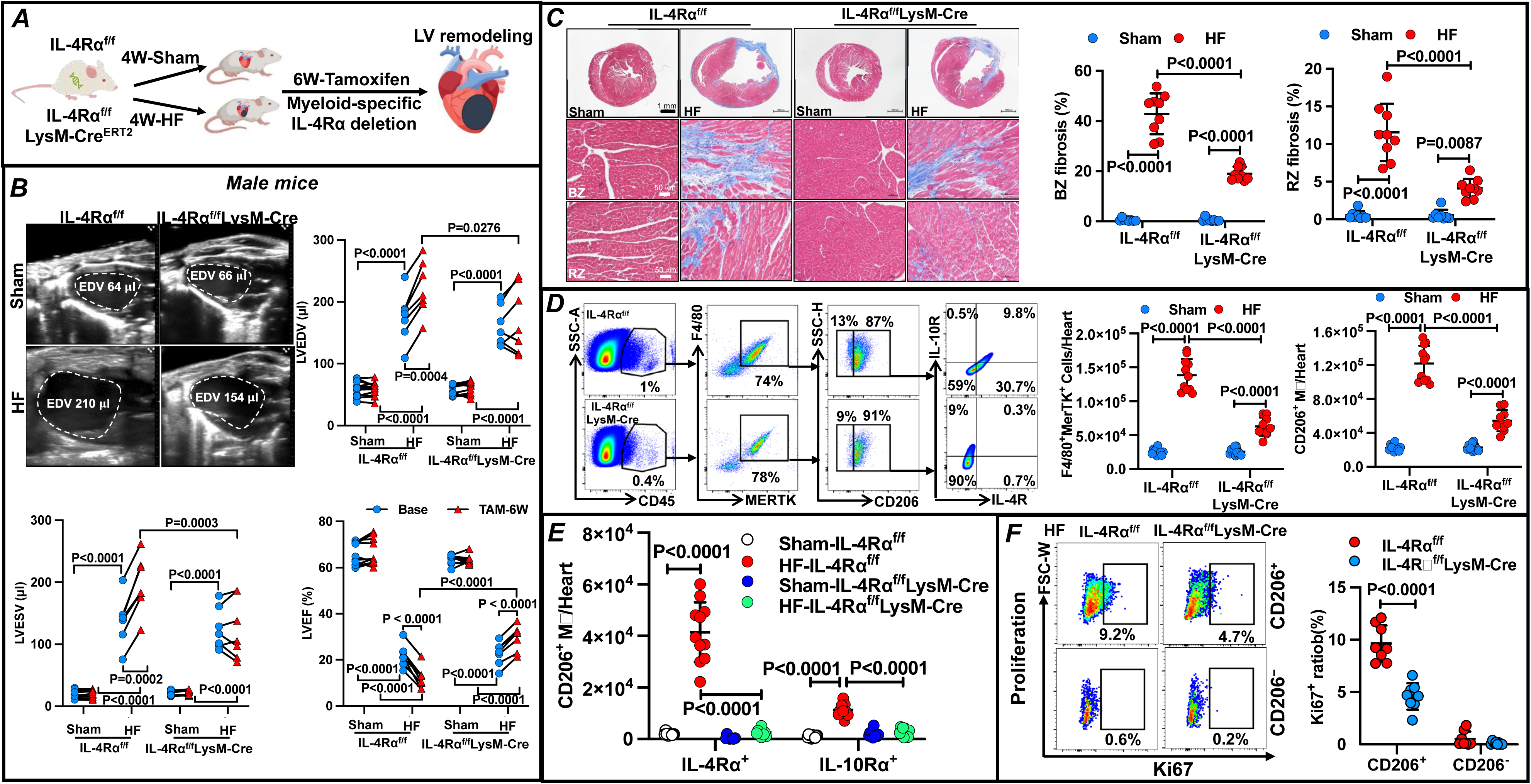
Myeloid-specific IL-4Rα deletion suppresses cardiac CD206^+^ macrophages and alleviates LV remodeling in heart failure (HF). *A*, Protocol for myeloid-specific IL-4Rα ablation in IL-4Rα^f/f^LysM-Cre^ERT2^ sham and HF mice (IL-4Rα^f/f^ mouse control) with tamoxifen administration in diet from 4 to 10 w post-MI or sham surgery. *B*, Representative long-axis 2D echocardiograms at end-diastole from male sham and HF mice after 6 w of tamoxifen (10 w post-surgery), along with directional changes in LV end-diastolic and end-systolic volume (EDV and ESV) and LV ejection fraction (EF) over this time. *C*, Representative trichrome stains of hearts from IL-4Rα^f/f^ and IL-4Rα^f/f^LysM-Cre^ERT2^ sham and HF mice after 6 w of tamoxifen (10 w post-surgery) along with fibrosis quantitation in the border and remote zone (BZ and RZ). *D* and *E*, Representative flow plots for cardiac macrophages (F4/80^+^MerTK^+^), CD206^+^ macrophages, and IL-4Rα and IL-10Rα expressing CD206^+^ subpopulations in IL-4Rα^f/f^ and IL-4Rα^f/f^LysM-Cre^ERT2^ HF mice, along with quantitative group data from both sham and HF male mice after 6 w of tamoxifen. *F*, Representative flow plots and quantitation of Ki67 expression in cardiac CD206^+^ and CD206^−^ macrophages from IL-4Rα^f/f^ and IL-4Rα^f/f^LysM-Cre^ERT2^ HF mice given tamoxifen for 6 w. Statistics: 2-way ANOVA with post-hoc Tukey’s multiple comparisons between groups in *B-E*; unpaired Student’s t test in in *F*. N=7-11 per group.

Comparable tamoxifen-induced effects were observed in female IL-4Rα^f/f^LysM-Cre^ERT2^ versus IL-4Rα^f/f^ HF mice with regard to improvements in LV remodeling and systolic function (albeit with less pronounced differences in LVEDV) (*Figure S9A-B*), heart and lung gravimetry and weight (*Figure S9C-D*), and reductions in cardiac leukocytes and total macrophages (Figure S10A), CD206^+^ and CD206^−^ macrophages, CD206^+^IL4Rα^+^ and CD206^+^IL10Rα^+^ macrophages, and CD206^−^IL4Rα^+^ macrophages (*Figures S10B-C*). As in males no effects of myeloid IL-4Rα deletion on any of these parameters were observed in sham-operated female mice. Collectively, these results indicate that CD206^+^IL4Rα^+^ macrophages are a key expanded myeloid cell population in the failing heart that is necessary for driving progressive inflammation and pathological cardiac remodeling, fibrosis, and dysfunction. Moreover, IL-4Rα expression marks a macrophage subpopulation with high proliferative capacity in the failing heart.

### IL-4Rα silencing *in vivo* reverses LV remodeling, reduces cardiac fibrosis, and improves neovascularization in HF

To explore whether the macrophage IL-4Rα axis could be exploited for therapeutic benefit in HF, we selectively silenced IL-4Rα expression *in vivo* using optimized antisense oligonucleotides (ASOs) targeting IL-4Rα mRNA.^40,41^ IL-4Rα signaling mainly occurs in hematopoietic and lymphoid cells.^45^ In humans, there is very low IL-4R RNA expression in heart cardiomyocytes and fibroblasts, with much more pronounced expression in immune cells, and in endothelial and smooth muscle cells (https://www.proteinatlas.org/ENSG00000077238-IL4R/single+cell+type/heart+muscle). To assure targeting of cardiac macrophages with this systemic approach, we performed flow cytometry to evaluate IL-4Rα expression in cardiac leukocytes. As shown in *Figure S11*, in both sham and failing hearts, ∼92% of all IL-4Rα^+^ CD45^+^ leukocytes were macrophages (F4/80^+^MERTK^+^), while ∼98% of IL-4Rα^+^ macrophages in failing hearts were CD206^+^. Moreover, circulating monocytes and neutrophils exhibited low levels of IL-4Rα expression (*Figure S2*). Hence, IL-4Rα ASOs represent a feasible and directed approach for dual inhibition of IL-4 and IL-13 signaling^45^ in cardiac CD206^+^IL-4Rα^+^ macrophages.

Male sham-operated and HF mice with comparable degrees of LV remodeling at 4 w post-MI were randomized to receive either IL-4Rα ASOs or MM oligos (40 mg/kg every 3 days) for 4 weeks (*Figure 6A*). Body weight was comparable between the two sham groups and two HF groups over the 4 w treatment period (*Figure S12A*). Serial echocardiography revealed significant and expected LV dilation and systolic dysfunction in both HF groups as compared to sham groups (*Figures 6B and S12B*). Sham-operated mice exhibited no changes in LVEDV, LVESV, stroke volume (SV), and LVEF during treatment with either IL-4Rα ASOs or MM oligos. HF mice treated with MM oligos evidence progressive increases in LV chamber size, and reductions in LVEF and SV, over 4 w. In contrast, HF-IL-4Rα ASO mice, which exhibited LV remodeling similar to HF-MM oligo mice at 4 w post-MI, exhibited significant reductions in EDV and ESV, and improvements in SV and EF, indicative of reverse remodeling. Lung and heart gravimetric analysis at 8 w post MI revealed substantially attenuated pulmonary edema and cardiac hypertrophy in HF-IL-4Rα ASO HF mice versus HF-MM oligo mice, without impact on spleen weight (*Figures 6C and S12C*).

**Figure 6.**
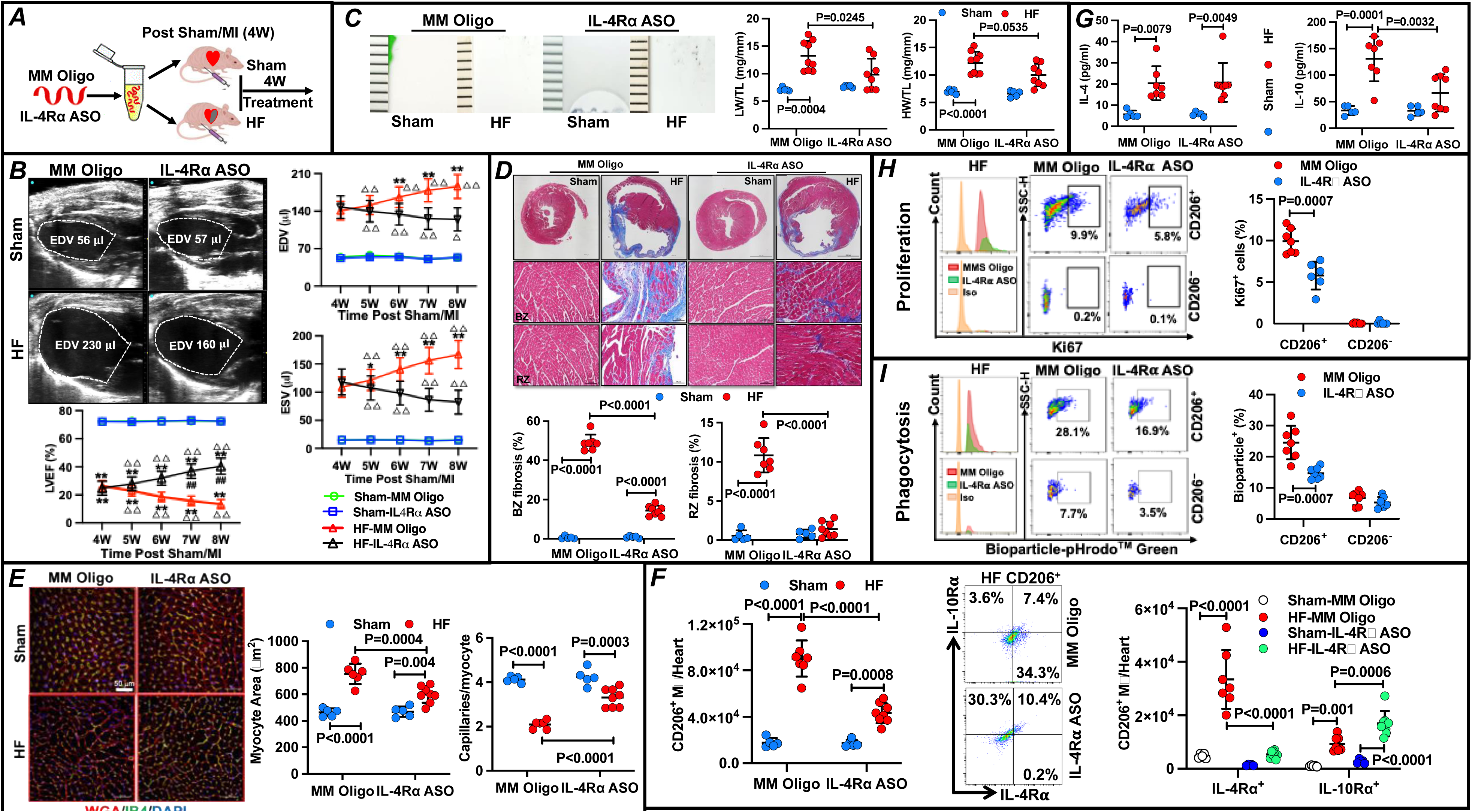
*In vivo* IL-4Rα silencing depletes cardiac CD206^+^ macrophages and reverses LV remodeling in heart failure (HF). ***A***, Protocol for treatment with IL-4Rα antisense oligonucleotides (ASOs) or mismatched (MM) control oligonucleotides (oligo) in HF mice from 4 to 8 w after non-reperfused myocardial infarction (MI) (sham controls). ***B***, Representative long-axis 2-dimensional echocardiograms at end-diastole from sham and HF mice treated with IL-4Rα ASOs or MM Oligos for 4 w (8 w post-surgery timepoint). Also shown are group data for LV end-diastolic and end-systolic volume (EDV and ESV) and ejection fraction (EF) over the treatment period. ***C***, Gross images of hearts from IL-4Rα ASO- and MM Oligo-treated sham and HF mice, and group data for lung and heart weight (LW and HW) normalized to tibia length (TL). ***D*,** Representative trichrome stains of IL-4Rα ASO- and MM Oligo-treated sham and failing hearts, and fibrosis quantitation in the border and remote zone (BZ and RZ). ***E***, Representative cardiac isolectin B4 (IB4, green) and WGA (red) staining and quantitation of myocyte cross-sectional area and capillary:myocyte ratio in the LV RZ from IL-4Rα ASO- and MM Oligo-treated sham and HF mice. ***F***, *Left*, quantitation of cardiac CD206^+^ macrophages, and *Middle* and *Right*, flow plots and quantitation of IL-4Rα and IL-10Rα expressing subpopulations, in sham and HF mice treated with IL-4Rα ASOs or MM Oligos for 4 w. ***G***, Serum levels of IL-4 and IL-10 from the same groups. ***H*** and ***I***, Representative flow cytometry histograms, dot plots, and quantitation of cardiac Ki67^+^ (***H***) or bioparticle^+^ (***I***) CD206^+^ and CD206^−^ macrophages in IL-4Rα ASO- and MM Oligo-treated HF mice. Statistics: repeated-measures 1-way ANOVA with post-hoc Dunnett’s multiple comparison test for time series within groups in ***B***; 2-way ANOVA with post-hoc Tukey’s multiple comparisons between groups in ***B-F, H, I***; unpaired Student’s t test in ***G***. \*\**p*<0.01 vs. respective Sham-MM Oligo; ^##^*p*<0.01 vs. HF-MM Oligo at the same week post-surgery; **^Δ^***p*<0.05, **^ΔΔ^***p*<0.01 vs. respective 4 w timepoint. N=5-8 per group.

Histological evaluation with trichrome staining indicated significantly increased border and remote zone interstitial fibrosis in MM oligo-treated failing hearts as compared to MM oligo-treated sham hearts (*Figure 6D*). Impressively, IL-4Rα ASO treatment profoundly decreased both border and remote zone fibrosis in failing hearts. Fibrotic area in IL-4Rα ASO-treated sham hearts was comparable to that of MM oligo-treated sham hearts. Isolectin B4 and wheat-germ agglutinin staining (*Figure 6E*) indicated that both HF-MM oligo mice and HF-IL-4Rα ASO mice exhibited significantly increased myocyte cross-sectional area and lower myocardial capillary:myocyte ratio versus their respective sham groups. Importantly, however, myocyte area was lower and capillary:myocyte ratio was higher in HF-IL-4Rα ASO mice versus HF-MM oligo mice, indicating alleviation of pathological hypertrophy and capillary rarefaction.

### IL-4Rα silencing in HF depletes cardiac CD206^+^IL-4Rα^+^ macrophages and suppresses their proliferative and phagocytic capacity

As shown in *Figures S13A and 6F*, 4 w of IL-4Rα ASO treatment significantly reduced the number of total CD45^+^ leukocytes (by ∼45%), total F4/80^+^MERTK^+^ macrophages (by∼53%), and both CD206^+^ and CD206^−^ macrophages (by ∼50%) in the failing heart. Moreover, IL-4Rα ASO treatment specifically and profoundly depleted CD206^+^IL-4Rα^+^ cardiac macrophages, while mildly augmenting CD206^+^IL-10Rα^+^ macrophages in the failing heart (*Figure 6F*). IL-4Rα ASO administration did not impact leukocyte and total macrophage abundance in sham hearts. Notably, IL-4Rα ASO therapy did not impact in circulating IL-4 levels (nor IL-13 and TGF-β1 levels) in HF mice, whereas serum IL-10 was significantly decreased (*Figures 6G and S13B*). Remarkably, total CD206**^−^** macrophages, including CD206**^−^**IL-4Rα^+^ and CD206**^−^**IL-10Rα^+^ macrophages, in failing hearts were also diminished by IL-4Rα ASO treatment (*Figure S13A*). Additionally, IL-4Rα ASO treatment impacted CD206^+^ macrophage function in failing hearts, significantly reducing (by ∼50%) both CD206^+^ macrophage cell proliferation (Ki67 staining) and phagocytic capacity (assessed in vivo using fluorescent bioparticles) in HF-IL-4Rα ASO mice as compared with HF-MM oligo mice (*Figures 6H-I*). The latter data are consistent with IL-4 serving as a potent stimulator of macrophage proliferation.^15,55^

IL-4Rα ASO treatment in HF mice also abrogated systemic inflammation as evidenced by normalization of circulating Ly6C^hi^ monocytosis (and levels of Ly6C^hi^IL-4Rα^+^ monocytes) and serum levels of pro-inflammatory protein mediators TNF-α, MCP-1, MIP-2α and IL-1β toward those observed in sham mice (*Figure S13C-D*). Serum IFN-γ levels, however, were not impacted by IL-4Rα ASO treatment and remained high in HF-IL-4Rα ASO mice as compared to sham. Importantly, IL-4Rα ASO treatment in HF mice also alleviated T-cell expansion in both the circulation and failing heart, with significant reductions in peripheral blood and cardiac CD4^+^ and CD8^+^ T cells in in HF-IL-4Rα ASO mice as compared with HF-MM oligo mice (*Figure S14*). Additionally, IL-4Rα expression was very low (∼1-2%) in blood and heart CD4^+^ and CD8^+^ T cells in both sham and HF mice treated with MM oligo or IL-4Rα ASOs. Taken together, these data indicate that in vivo IL-4Rα mRNA silencing was highly effective in improving pathological cardiac remodeling, hypertrophy, fibrosis, and dysfunction, and in suppressing local and systemic inflammation in chronic ischemic HF.

### Fizz1 is essential for CD206^+^ macrophage-induced myofibroblast activation

FIZZ1, also known as resistin-like molecule α (*Retnla*), is a 9.4 kDa cysteine-rich secreted protein produced mostly by macrophages during Th2 responses.^50–52^ As described above, macrophage Fizz1 expression exponentially increased exclusively after M[IL-4] polarization (>7,500-fold after 24 h and >100,000-fold after 48 h) polarization and not with M[IL-10] polarization (*Figures 3B and S5B*), and coincided with M[IL-4] macrophage-induced myofibroblast differentiation of cMSCs (*Figure 3C*). Accordingly, we further evaluated the role of Fizz1 in pro-fibrotic cellular responses. Exposure of cultured cMSCs to recombinant mouse (rm) Fizz1 induced, in a dose-dependent manner, the expression of proliferative genes *PCNA, Mki67, CyclinB1* and *CyclinD1* early (2 h), and profibrotic genes *α-SMA, Col1α1, Col1α2*, and *Col3α1* late (16 h), after treatment (*Figure S15*), analogous to changes observed with M[IL-4] medium treatment (*Figure 3C*). To determine the need for FIZZ1 in M[IL-4]-induced cMSC responses, we used anti-Fizz1 antibody to neutralize endogenous Fizz1 in M[IL-4] conditioned medium. As shown in *Figure S16*, compared to IgG control, anti-Fizz1 significantly reduced the expression of mitotic genes, *α-SMA* (by 40%) and *Col3α1* in M[IL-4] conditioned media-treated cMSCs. Furthermore, anti-Fizz1 abrogated collagen gel contraction induced by M[IL-4] conditioned media (*Figure 7A*), a phenomenon recapitulated by the use of Fizz1^-/-^ BMDMs (*Figure 7B),* indicating that secreted Fizz1 is essential for M[IL-4]-induced cMSC myofibroblast differentiation and activation.

**Figure 7.**
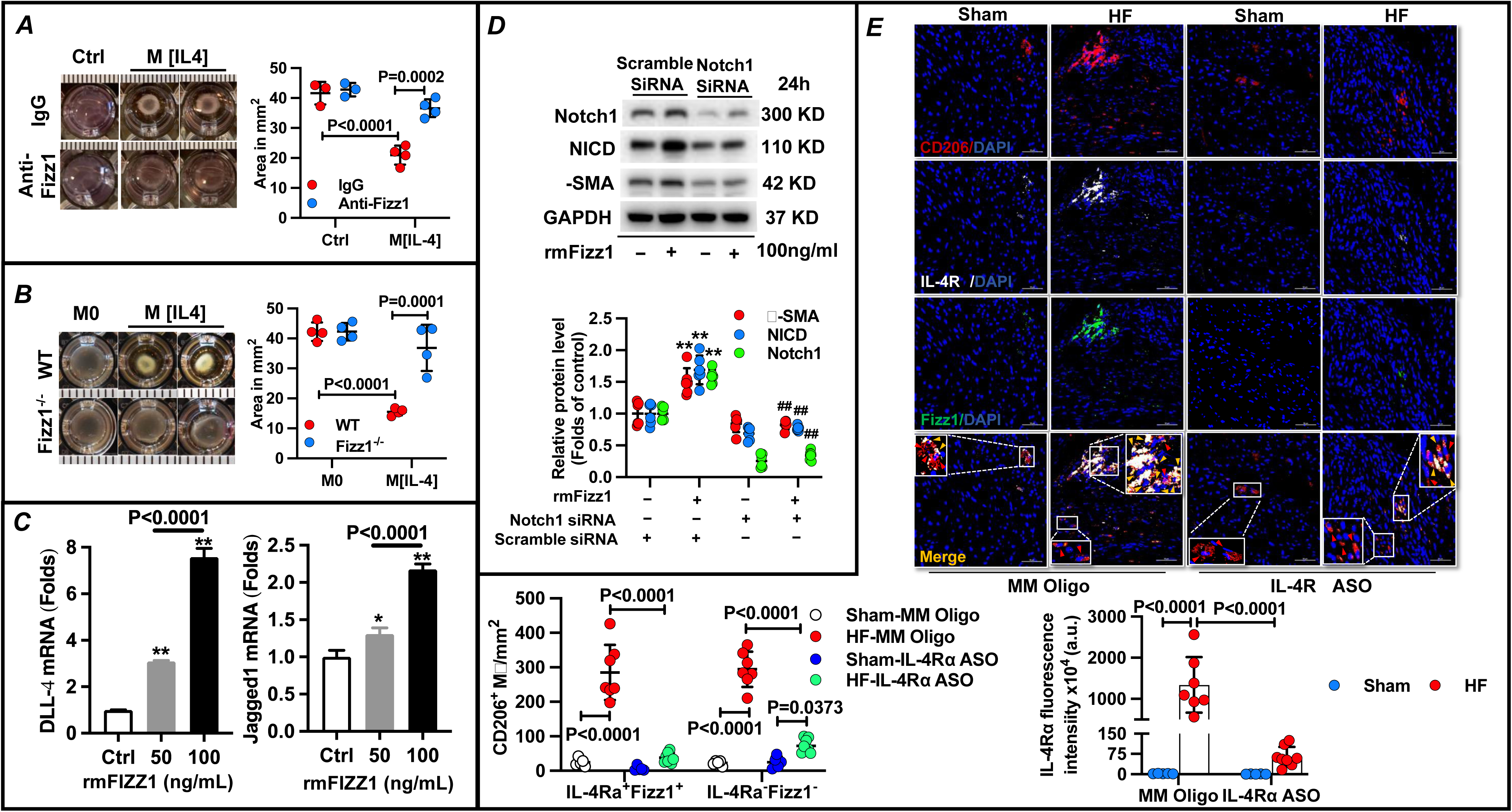
Fizz1 is essential for CD206^+^ macrophage-induced myofibroblast differentiation of cardiac mesenchymal stem cells (cMSCs). *A*, cMSC-imbued collagen gel contraction assays upon exposure to M[IL-4] bone marrow-derived macrophage (BMDM) conditioned media or media alone control (Ctrl) containing isotype IgG or anti-Fizz1 antibody with corresponding quantitation of gel areas. *B*, cMSC gel contraction assays upon exposure to wild-type (WT) and Fizz1^-/-^ M0 or M[IL-4] BMDM conditioned media, and quantitation of gel areas. N=3-5 per group for *A-B*. *C*, cMSC DLL-4 and Jagged1 gene expression after 30 minutes of treatment with either vehicle (Ctrl) or recombinant mouse (rm) Fizz1 50 ng/ml and 100 ng/ml. *D*, Immunoblotting and group quantitation for Notch1, Notch intracellular domain (NICD), and α-smooth muscle actin (α-SMA) protein expression in cMSCs treated with either vehicle Ctrl or rmFizz1 for 24 h following scrambled or Notch1 siRNA transfection. GAPDH loading control. *E*, Representative low and high magnification confocal images of immunostains for CD206 (red), IL-4Rα (white) and Fizz1 (green) of hearts from sham and HF hearts (border zone) from MM Oligo- or IL-4Rα ASO-treated mice, with quantitation of CD206^+^IL-4Rα^+^Fizz1^+^ and CD206^+^IL-4Rα^−^Fizz1^−^ macrophages, and IL-4Rα fluorescent intensity (arbitrary units [a.u.]) n=5-8/group. DAPI (blue) was used to label nuclei. Red arrows indicate CD206^+^IL-4Rα^−^Fizz1^−^ cells and yellow arrows indicate CD206^+^IL-4Rα^+^Fizz1^+^ cells. Statistics: 2-way ANOVA followed by Tukey’s multiple comparison test, except 1-way ANOVA followed by Tukey’s multiple comparison test in panel *C*; \**p*<0.05, \*\**p*<0.01 vs. Ctrl in *C*; \*\**p*<0.01 vs. Ctrl-scramble siRNA, ^##^*p*<0.01 vs. rmFIZZ1-scramble siRNA in *D*. N=6 per group. MM, mismatched; ASO, antisense oligonucleotide.

Notch signaling is critical for cell proliferation, differentiation, and development, and homeostasis;^56^ Notch ligand binding to Notch receptors leads to release of Notch intracellular domain (NICD) and subsequent transcription of target genes, including *α-SMA, Cyclin D1, Cyclin A* and NF-κB.^56^ As α-SMA expression is critical for myofibroblast fate of mesenchymal stromal cells,^33,57^, we assessed the importance of Notch signaling in Fizz1-mediated α-SMA expression in cMSCs. Fizz1 exposure significantly upregulated cMSC gene expression of the Notch1 ligands DLL-4 and Jagged1 within 30 minutes (*Figure 7C*), followed by increased protein expression of Notch1, NICD and α-SMA within 24 h (*Figure 7D*). Importantly, Notch1 siRNA knockdown prevented α-SMA (and NICD) upregulation in Fizz1-treated cMSCs, suggesting a critical role for DLL-4/Jagged-Notch1 signaling in the pro-fibrotic effects of M[IL-4] macrophage-secreted FIZZ1.

In view of the importance of FIZZ1 in M[IL-4]-induced myofibroblast differentiation, we next examined Fizz1 expression and spatial distribution using immunostaining in failing hearts from MM oligo- and IL-4Rα ASO-treated mice (8 w post-MI). Consistent with flow data, a robust (∼11-fold) expansion of CD206^+^ macrophages (both IL-4Rα^+^ and IL-4Rα^-^) was observed in MM oligo-treated failing hearts versus sham hearts, especially in the border zone (*Figure 7E*). IL-4Rα staining was observed near exclusively in CD206^+^ cells (*Figure S17*), further supporting CD206^+^ macrophage-specificity of IL-4Rα silencing, and Fizz1^+^ staining was near exclusively spatially localized to IL-4Rα^+^ cells (∼98% Fizz1^+^) in all groups (*Figure S17*). IL-4Rα^+^ Fizz1^+^ cells comprised ∼50% cardiac CD206^+^ macrophages in HF-MM oligo mice (*Figure 7E*). IL-4Rα ASO treatment significantly decreased IL-4Rα^+^Fizz1^+^ (and IL-4Rα**^−^**Fizz1**^−^**) CD206^+^ macrophages in failing hearts, and also diminished IL-4Rα expression (assessed by fluorescence intensity) (*Figure 7E*). (*Figures 7E and Figure S17*). Hence, IL-4Rα silencing in HF reduced CD206^+^ macrophage FIZZ1 expression in the failing heart concomitantly with improvements in cardiac fibrosis. Taken together with the *in vitro* studies above, our results suggest that *Fizz1* may be a key contributor to the tissue level pro-fibrotic effects of CD206^+^IL-4Rα^+^ macrophages in the failing heart.

### IL-4Rα^+^Fizz3^+^ alternatively activated macrophages expand and directly correlate with fibrosis in human failing hearts

To understand translational relevance, we lastly explored whether alternatively activated resident macrophages are expanded in human HF. As mentioned above, resident cardiac macrophages express classical M2-like markers *Mrc1* (*CD206*) and *CD163*.^2,6,13,14^ Cardiac mononuclear cells were isolated from the LV apex procured from advanced HF patients during LVAD implantation, and from control donor hearts unsuitable for transplantation. Analogous to murine HF, flow cytometry indicated significant expansion (∼10-fold) of CD206^+^ macrophages (CD45^+^Autofluorescence^+^CD64^+^) in human failing hearts versus control, with ∼20% of CD206^+^ macrophages expressing CCR2 (*Figure 8A*). Human failing hearts also exhibited substantial (>200-fold) interstitial fibrosis versus control hearts, with most (∼95%) CD163^+^ macrophages found in association with areas of collagen turnover (*Figure 8B*), analogous to murine failing hearts. *Fizz3* (resistin, *Retn*), the human homolog of murine *Fizz1,* is expressed by M[IL-4/13] human macrophages and resembles mouse FIZZ1 with respect to sequence and function.^58,59^ Accordingly, we performed immunostaining for IL-4Rα, Fizz3 and CD163, a widely used marker for alternatively-activated human macrophages.^60^ As shown in *Figure 8C*, human failing hearts exhibited significant expansion (∼6-fold) of CD163^+^ macrophages (both IL-4Rα^+^ and IL-4Rα^-^) as compared to control hearts. Fizz3^+^ staining was near exclusively spatially localized to IL-4Rα^+^ cells (∼98% Fizz3^+^) in both human control and failing hearts, similar to the IL-4Rα and Fizz1 expression pattern observed in mouse hearts (*Figure S17*). Importantly, IL-4Rα^+^Fizz3^+^ cells comprised the predominant CD163^+^ macrophage subset in human failing hearts, with approximately two-thirds of CD163^+^ macrophages expressing IL-4Rα and Fizz3^+^ (*Figure 8C*). CD163^+^ macrophage abundance directly correlated with cardiac fibrosis in human HF. Hence, analogous to murine HF, human failing hearts also evidence expansion of alternatively activated macrophages, likely with augmented IL-4 signaling and contributing to pro-fibrotic responses.

**Figure 8.**
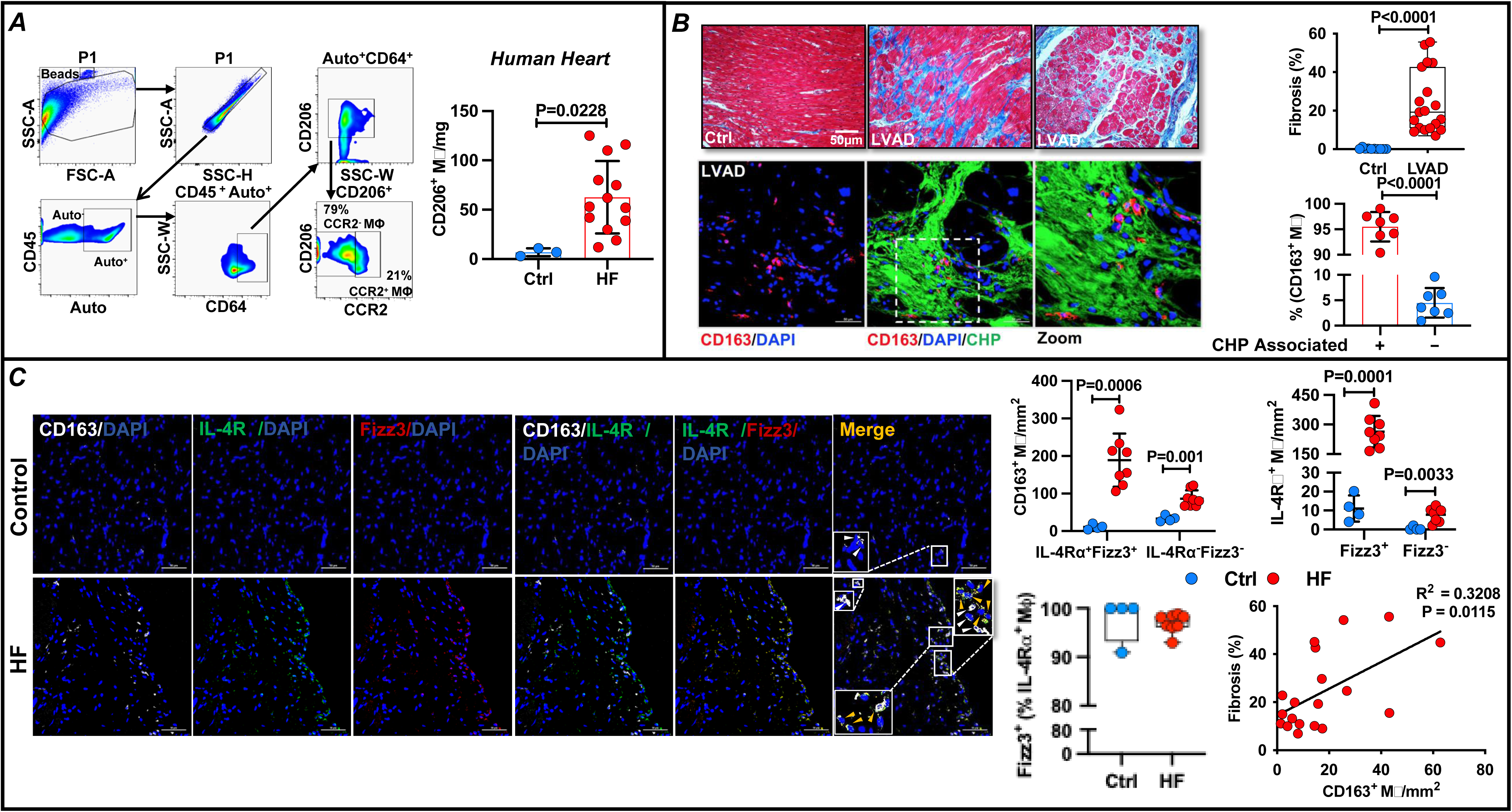
Alternatively activated cardiac macrophages expand, express IL-4Rα and FIZZ3, and directly correlate with fibrosis in human heart failure (HF). *A*, Representative flow plots for CD45^+^Autofluorescence(Auto)^+^CD64^+^CD206^+^ macrophages and their CCR2 expression, and group quantitation for CD206^+^ macrophages in normal control (Ctrl; n=3) and failing (n=13) human hearts (from LV assist device core tissue). *B*, Representative trichrome stains (*Top*) and low and high magnification (immuno)stains for CD163 (red) and collagen-hybridizing peptide (CHP, green) (*Bottom*) of Ctrl and failing human hearts, with corresponding quantitation of cardiac fibrosis (10 Ctrl, 19 failing hearts) and CHP-associated and - unassociated CD163^+^ macrophage frequency (n=7/group). *C*, Low and high magnification confocal images of immunostains for CD163 (white), IL-4Rα (red) and Fizz3 (green) in normal (Ctrl; n=4) and failing (n=8) human hearts (HF), and quantitation of IL-4Rα^+^Fizz3^+^ and IL-4Rα^−^Fizz3^−^ CD163^+^ macrophages. DAPI (blue) was used to label nuclei. White arrows indicate CD163^+^IL-4Rα^−^Fizz3^−^ cells, and yellow arrows indicate CD163^+^IL-4Rα^+^Fizz3^+^ cells. Also shown is the correlation between CD163^+^ macrophage abundance and LV fibrosis in human HF, n=19. Statistics: Mann-Whitney test in *A*; unpaired Student’s t test in *B-C;* Pearson R correlation test in *C, lower right panel*.

## DISCUSSION

We describe a novel role for cardiac CD206^+^ macrophages alternatively activated by IL-4 receptor signaling in the progression of LV remodeling in ischemic cardiomyopathy. There are several key findings. First, macrophages expressing CD206 comprised most macrophages in the naïve heart. Following MI, cardiac CD206^+^ macrophages progressively expanded, especially during the later stages of LV remodeling such that they comprised the predominant macrophage population in chronic HF. Second, CD206^+^ macrophages were exclusively proliferative and primarily CCR2^-^, MHCII^hi^, and LYVE1^+^ suggesting a resident cell population, and correlated with LV dysfunction and fibrosis. Second, a sizable subpopulation of these macrophages in HF expressed IL-4Rα; this occurred with attendant increased systemic levels of IL-4. Third, M[IL-4] (but not M[IL-10]) CD206^+^ BMDMs induced remarkable upregulation of *Fizz1,* and robust cMSC myofibroblast differentiation. M[IL-4] CD206^+^ macrophage-induced myofibroblast differentiation and activation were dependent on FIZZ1 secretion and mediated in part by FIZZ1 induction of DLL-4/Jagged1-Notch1 signaling in cMSCs. Fourth, intramyocardial adoptive transfer of M[IL-4] macrophages was sufficient to induce pathological cardiac remodeling and fibrosis. Fifth, myeloid-specific IL-4Rα deletion tempered cardiac CD206^+^ macrophage proliferation and alleviated LV dysfunction, fibrosis, and adverse remodeling. Sixth, IL-4Rα silencing *in vivo* was highly effective in depleting CD206^+^IL-4Rα^+^ macrophages, reducing FIZZ1 expression in the failing heart, and reversing LV remodeling in mice with established HF. Lastly, humans exhibit expansion of alternatively activated macrophages in the failing heart marked by CD206 and CD163 expression, with predominance of a subpopulation expressing IL-4Rα and Fizz3, the human homolog of *Fizz1*. Collectively, these data establish that an expanded pool of alternatively activated CD206^+^IL-4Rα^+^ cardiac macrophages are key drivers of LV remodeling in HF, inducing a Th2/M2-like immuno-fibrotic response in part dependent on FIZZ1. Targeting IL-4 signaling in CD206^+^ macrophages may represent a fruitful therapeutic approach to limit long-term cardiac remodeling in HF.

*Mrc1* (*Cd206*) and *Cd163* are typically expressed by resident cardiac macrophages under steady state conditions.^2,6,13,14^ Moreover, in the naïve heart, CD206^+^ macrophages express high levels of classical M2-like genes such as *Ym1* and *Fizz1* and are primarily CCR2^-^,^15^ suggesting a macrophage subset with minimal monocyte-dependence.^6^ Prior work has shown that CD206^+^ macrophages expand during the first week after acute MI in the infarct and border zone to promote infarct repair and functional restoration.^15–17^ Administration of IL-4 acutely (but not late) after MI also expands CD206^+^ macrophages in the infarcted heart and recapitulates beneficial effects on post-MI cardiac healing.^15,24^ However, the role of this macrophage population, and the impact of macrophage IL-4 signaling, in chronic LV remodeling and ischemic HF has not been defined. The chronically failing heart is typically marked by excessive tissue fibrosis.^61^ Notably, in chronic pulmonary,^62,63^ hepatic,^41^ and renal^25^ disease models marked by excessive fibrosis, alternatively activated (CD206^+^ or CD163^+^) macrophage activation mediated by Th2 cytokines such as IL-4 contribute to disease progression. While we have previously shown that the post-MI failing heart exhibits increased IL-4 expression and a predominant Th2 CD4^+^ T-cell profile,^26^ whether a similar pathophysiological axis contributes to LV remodeling in ischemic cardiomyopathy is unknown.

Our studies show that CD206^+^ macrophages progressively expand after MI, with particularly robust levels during the later stages of LV remodeling. Beyond the initial infiltration of predominantly (>80%) CD206^−^ macrophages into the heart early (48 h) after MI, CD206^+^ macrophages were the main subtype in the remodeling heart at 1 w post-MI and beyond, comprising ∼85% of all macrophages in HF. Moreover, in the failing heart 8 w post-MI, nearly half of all cardiac CD206^+^ macrophages expressed IL-4Rα. Notably, cardiac macrophage proliferation was observed exclusively in CD206^+^ subpopulation in the failing heart. Although spatially expanded throughout the myocardium, CD206^+^ macrophages were most abundant in the infarct scar and border zone, and primarily in regions of collagen turnover and fibrosis. Heightened macrophage proliferation was also mainly seen in the scar and border zones. This suggests that progressive CD206^+^ macrophage expansion in these regions may contribute to extracellular matrix remodeling, infarct expansion and LV chamber dilatation over time. Human HF also evidenced marked expansion of alternatively activated (CD206^+^ or CD163^+^) cardiac macrophages (consistent with a prior study of macrophages isolated from human failing hearts^64^) in association with regions of collagen denaturation and turnover, with a predominance of cells expressing IL-4Rα analogous to murine HF. In this regard, IL-4 is a known stimulator of tissue macrophage proliferation and resident macrophage self-renewal^55,65^ that also augments macrophage CD206 expression^66^ and promotes M2-like cardiac macrophage differentiation.^15^ Consistent with our prior studies,^26^ IL-4 levels were increased both locally in the heart and systemically, suggesting that heightened IL-4 signaling may contribute importantly to sustaining local inflammation and macrophage expansion in failing myocardium.

In both murine and human HF, the expanded cardiac CD206^+^ macrophage population was primarily CCR2^−^ (∼80%), MHCII^hi^, and LYVE1^+^. In naïve^15^ and in sham-operated hearts, CD206^+^ macrophages were also primarily CCR2^−^, albeit to much greater degrees (∼95%) than in failing hearts. As LYVE1 is considered a marker of resident macrophages,^6^ this suggests a predominantly resident macrophage population that is expanded in failing hearts, but also with a dual (albeit lesser) contribution from CCR2^+^ monocyte-derived cells. Prior work has shown that after MI the absence of CCR2 expression does not exclude a monocyte origin as recruited macrophages over time may exhibit transcriptional identities nearly identical to resident macrophages.^6^ Our study design did not incorporate macrophage lineage tracing to definitively address this question. Nonetheless, our data are consistent with prior work indicating that cardiac macrophages derive from both local macrophage proliferation and recruited monocytes (locally-sourced cells comprising ∼75%) in chronic ischemic HF.^3,5^ Given our observations that the proportion of CCR2^+^ cells increased significantly in failing hearts, and that cardiac macrophage abundance in post-MI HF was paralleled by similar dynamic changes in Ly6C^high^ blood monocytes that source infiltrating macrophages,^3,5,6,10^ we propose that the expanded and proliferative cardiac CD206^+^macrophage population derives from both resident and recruited cells.

We evaluated the pathophysiological importance of CD206^+^IL-4Rα^+^ expansion in the failing heart by experimentally manipulating IL-4Rα signaling, which occurs largely in immune cells.^45^ While both M[IL-4] and M[IL-10] macrophage polarization induced the upregulation of signature alternative macrophage activation genes, M[IL-4] polarization exclusively resulted in extraordinary upregulation of *Fizz1*, and functional myofibroblast differentiation of cMSCs, together with enhanced cMSC proliferative gene expression. Importantly, intramyocardial adoptive transfer of M[IL-4], but not M[IL-10], macrophages produced significant long-term LV chamber dilatation, dysfunction, and fibrosis in naïve recipients. Hence, IL-4-mediated activation of macrophages was sufficient to recapitulate chronic cardiac remodeling.

To establish the necessity of CD206^+^IL-4Rα^+^ macrophages in the pathogenesis of chronic LV remodeling, we evaluated the effects of myeloid IL-4Rα deletion during established HF on subsequent LV remodeling. This manipulation primarily impacted macrophages in HF, given the very low levels of IL-4Rα in neutrophils and monocytes. Notably, loss of macrophage IL-4 signaling via IL-4Rα suppressed CD206^+^ macrophage proliferation, abrogated the progression of LV remodeling, improved LV systolic function and fibrosis, while suppressing both CD206^+^ and CD206^−^ macrophages in the heart as well as systemic Ly6C^hi^ monocytosis in the blood. Beneficial effects of macrophage IL-4Rα deletion were observed in both male and female mice. Hence, CD206^+^IL4Rα^+^ macrophages are indispensable for sustaining a chronic para-inflammatory response in failing myocardium that drives progressive cardiac remodeling in HF.

To explore a potential therapeutic avenue, we performed *in vivo* gene silencing of IL-4Rα using ASOs, an approach that primarily targeted cardiac CD206^+^ macrophages given the aforementioned low distribution of IL-4Rα expression observed in other myeloid and lymphoid cell populations in chronic HF. IL-4Rα silencing in chronic ischemic HF very effectively depleted cardiac CD206^+^IL-4Rα^+^ macrophages and suppressed their proliferation and phagocytic capacity, while inducing impressive reverse LV remodeling with improvements in cardiac fibrosis, neovascularization, and function, and suppression of both local and systemic inflammation (*e.g*., blood and cardiac CD4^+^ and CD8^+^ T cell levels). Taken together, our results underscore the pathological effects of chronic macrophage IL-4Rα activation in the failing heart and establish CD206^+^IL-4Rα^+^ macrophages as key mediators of pathological remodeling of the failing heart. Our data support biological or pharmacological blockade of the IL-4/IL-4Rα pathway as a novel immunomodulatory paradigm in HF. For instance, antagonistic antibodies against IL-4Rα are currently being developed for allergic disorders in humans,^67^ and could potentially be repurposed for HF.

CD206^+^ macrophage abundance in murine failing hearts (or CD163^+^ macrophage abundance in human HF) correlated with the extent of cardiac fibrosis in chronic HF, and a prominent feature of IL-4-mediated CD206^+^ macrophage activation *in vitro* and *in vivo* was the induction of pro-fibrotic responses. Our data suggest that the protein FIZZ1 secreted by M[IL-4] macrophages may play an important role in this crosstalk, given: 1) *in vitro* studies demonstrating incredibly robust macrophage *Fizz1* upregulation upon IL-4 stimulation, 2) FIZZ1 dependence of the induction of cMSC myofibroblast differentiation by M[IL-4] macrophages, potentially via DLL-4/Jagged1-Notch1 signaling, 3) exclusive expression of Fizz1 by cardiac CD206^+^IL-4Rα^+^ macrophages in chronic HF, and 4) exclusive expression of FIZZ3 (resistin), the human homolog of FIZZ1,^58^ by CD163^+^IL-4Rα^+^ macrophages in human failing hearts. FIZZ1 is secreted during Th2-type immune responses in mice.^52,68^ and induces myofibroblast differentiation and fibrosis in the lung,^53,54^ whereas FIZZ3 induces LV fibrosis and dysfunction when overexpressed in rat heart^69^ and is a serum risk prediction biomarker in HF patients.^70^ Our data are consistent with a key pro-fibrotic function for FIZZ1 in the failing heart, secreted by alternatively-activated CD206^+^IL-4Rα^+^ macrophages as part of an overall Th2/M2-like inflammatory milieu in the failing heart.^26^ The elucidation of the precise role of Fizz1/Fizz3, and the potential for therapeutic Fizz1/Fizz3 blockade in HF, will require further study.

In summary, we have established a novel immunoinflammatory framework in the failing heart with an expanded pool of alternatively activated CD206^+^IL-4Rα^+^ macrophages as key drivers of LV remodeling. These alternatively-activated macrophages induce a para-inflammatory, immuno-fibrotic response, in part dependent on FIZZ1 expression, that promotes cardiac fibrosis and dysfunction. Our findings identify IL-4 signaling in CD206^+^ macrophages, and potentially FIZZ1/FIZZ3 elaboration, as novel and appealing therapeutic targets for immunomodulation to limit long-term cardiac remodeling in ischemic cardiomyopathy and HF.

## Data Availability

All data are available upon reasonable request.

## ACKNOWLEDGEMENTS AND FUNDING

This work was supported by National Institutes of Health R01 HL157999 and HL147549 grants to SDP, and R01 HL137046 to TH.

## COMPETING INTERESTS

There are no relevant author financial interests.

## NONSTANDARD ABBREVIATIONS AND ACRONYMS

LV: left ventricular

MI: myocardial infarction

HF: heart failure

EF: ejection fraction

EDV: end-diastolic volume

ESV: end-systolic volume

LVAD: left ventricular assist device

BMDM: bone marrow-derived macrophage

cMSC: cardiac mesenchymal stem cell

ASO: antisense oligonucleotide

CCR2: C-C motif chemokine receptor 2

IL: interleukin

TGF-β: tumor growth factor-β

Arg1: arginase 1

FIZZ: found in inflammatory zone

Col: collagen

PDGF: platelet-derived growth factor

α-SMA: α-smooth muscle actin

NICD: Notch intracellular domain

## Notes

### Competing Interest Statement

The authors have declared no competing interest.

### Author Declarations

University of Alabama at Birmingham (UAB) Institutional Review Board (IRB) approved the human studies.

